# Lowering of circulating sclerostin may increase risk of atherosclerosis and its risk factors: evidence from a genome-wide association meta-analysis followed by Mendelian randomization

**DOI:** 10.1101/2022.06.13.22275915

**Authors:** Jie Zheng, Eleanor Wheeler, Maik Pietzner, Till Andlauer, Michelle Yau, April E. Hartley, Ben Michael Brumpton, Humaira Rasheed, John P Kemp, Monika Frysz, Jamie Robinson, Sjur Reppe, Vid Prijatel, Kaare M Gautvik, Louise Falk, Winfried Maerz, Ingrid Gergei, Patricia A Peyser, Maryam Kavousi, Paul S. de Vries, Clint L. Miller, Maxime Bos, Sander W. van der Laan, Rajeev Malhotra, Markus Herrmann, Hubert Scharnagl, Marcus Kleber, George Dedoussis, Eleftheria Zeggini, Maria Nethander, Claes Ohlsson, Mattias Lorentzon, Nick Wareham, Claudia Langenberg, Michael V. Holmes, George Davey Smith, Jonathan H. Tobias

**Author notes:** Correspondence to: Jie Zheng, professor in Aetiological Epidemiology, Shanghai National Clinical Research Center for Endocrine and Metabolic Diseases, Key Laboratory for Endocrine and Metabolic Diseases of the National Health Commission of the PR China, Shanghai Institute of Endocrine and Metabolic Diseases, Department of Endocrine and Metabolic Diseases, Ruijin Hospital, Shanghai Jiao Tong University School of Medicine, Shanghai, China; and MRC Integrative Epidemiology Unit (IEU), Bristol Medical School, University of Bristol, Oakfield House, Oakfield Grove, Bristol, BS8 2BN, United Kingdom., George Davey Smith, professor in Clinical Epidemiology, MRC Integrative Epidemiology Unit (IEU), Bristol Medical School, University of Bristol, Oakfield House, Oakfield Grove, Bristol, BS8 2BN, United Kingdom. E-mail: kz.davey-, Jonathan H. Tobias, professor in Rheumatology, MRC Integrative Epidemiology Unit (IEU), Bristol Medical School, University of Bristol, Oakfield House, Oakfield Grove, Bristol, BS8 2BN, United Kingdom; and Musculoskeletal Research Unit, University of Bristol, Level 1 Learning and Research Building, Bristol, BS10 5NB, United Kingdom.

## Abstract

Sclerostin inhibition is a new therapeutic approach for increasing bone mineral density (BMD) but its cardiovascular safety is unclear. We conducted a genome-wide association study (GWAS) meta-analysis of circulating sclerostin in 33,961 Europeans followed by Mendelian randomization (MR) to estimate the causal effects of sclerostin on 15 atherosclerosis-related diseases and risk factors. GWAS meta-analysis identified 18 variants independently associated with sclerostin, which including a novel *cis* signal in the *SOST* region and three trans signals in B4GALNT3, RIN3 and SERPINA1 regions that were associated with opposite effects on circulating sclerostin and eBMD. MR combining these four SNPs suggested lower sclerostin increased hypertension risk (odds ratio [OR]=1.09, 95%CI=1.04 to 1.15), whereas bi-directional analyses revealed little evidence for an effect of genetic liability to hypertension on sclerostin levels. MR restricted to *cis (SOST)* SNPs additionally suggested sclerostin inhibition increased risk of type 2 diabetes (T2DM) (OR=1.26; 95%CI=1.08 to 1.48) and myocardial infarction (MI) (OR=1.31, 95% CI=1.183 to 1.45). Furthermore, these analyses suggested sclerostin inhibition increased coronary artery calcification (CAC) (β=0.74, 95%CI=0.33 to 1.15), levels of apoB (β=0.07; 95%CI=0.04 to 0.10; this result was driven by rs4793023) and triglycerides (β=0.18; 95%CI=0.13 to 0.24), and reduced HDL-C (β=-0.14; 95%CI=-0.17 to -0.10). This study provides genetic evidence to support a causal effect of sclerostin inhibition on increased hypertension risk. *Cis*-only analyses suggested that sclerostin inhibition additionally increases the risk of T2DM, MI, CAC, and an atherogenic lipid profile. Together, our findings reinforce the requirement for strategies to mitigate against adverse effects of sclerostin inhibitors like romosozumab on atherosclerosis and its related risk factors.

---

Inhibition of sclerostin is a therapeutic approach to increasing bone mineral density (BMD) and lowering fracture risk in patients with osteoporosis. However, two phase III trials of romosozumab, a first-in-class monoclonal antibody that inhibits sclerostin, reported higher numbers of cardiovascular serious adverse events in the romosozumab treated group over its comparator(*1*)(*2*). However, a similar imbalance of cardiovascular disease (CVD) was not seen in another study comparing romosozumab to placebo(*3*). Possibly, these different results reflect a beneficial effect of bisphosphonate treatment on risk of CVD. Another bisphosphonate, zoledronate, has been found to decrease all-cause mortality, to which reduced CVD mortality may contribute(*4*). However, a beneficial effect on mortality was not borne out in a meta-analysis of drug trials of zoledronate and other bisphosphonates(*5*). The role of sclerostin in the vasculature is unknown, though some studies have shown that its inhibition may promote vascular calcification, which would increase the risk of CVD(*6*). Given these concerns of CVD safety, marketing authorization for romosozumab indicates previous myocardial infarction (MI) or stroke as contraindications, underlying the urgent need to understand the causal role of sclerostin inhibition on CVD outcomes, so that further steps can be taken to mitigate these potential adverse effects.

Contrary to trial evidence suggesting an increase in CVD risk following sclerostin inhibition, our recent observational study found that sclerostin levels are positively associated with CVD severity and mortality, partly explained by a relationship between higher sclerostin levels and major CVD risk factors(*7*). Equivalent findings apply to BMD, with sclerostin levels found to be positively related to BMD(*8*), despite trial evidence suggesting that sclerostin lowering increases BMD(*1*)(*2*). Such discrepancies may reflect the influence of confounders or reverse causality on findings from observational studies(*9*). Mendelian randomization (MR) uses genetic variants as proxies for an exposure to estimate the causal effect of a modifiable risk factor on a disease(*10*)(*11*), in order to avoid bias from confounders or reverse causality. For example, in a recent MR study using BMD-associated variants in the *SOST* region as a proxy for sclerostin inhibition, Bovijn *et al.* found genetic evidence consistent with a potential adverse effect of sclerostin inhibition on CVD-related events(*12*). However, as discussed in the recent European Medicines Agency report on Romosozumab(*13*), this study has some weaknesses. For example, the *SOST* single nucleotide polymorphisms (SNPs) used in the analysis by Bovijn *et al.* are >30kb downstream of the target gene. Another MR study using sclerostin gene expression in arterial and heart tissue as the exposure was interpreted as showing no causal effect of sclerostin expression on risk of MI or stroke(*14*).

An alternative approach to instrument selection is to use SNPs identified from a well-powered genome-wide association study (GWAS) of circulating sclerostin. In an earlier GWAS of sclerostin levels, we identified three *trans*-acting genetic variants associated with sclerostin, including a top variant in the *B4GALNT3* region. However, we only observed marginal genetic associations in the *cis*-*SOST* region and had limited power to examine causal relationships with extra-skeletal phenotypes(*15*). Therefore, a GWAS of circulating sclerostin including more participants is needed to identify more reliable genetic predictors, including in the *cis*-acting region. A further consideration is that a bidirectional causal pathway appears to exist between sclerostin and BMD, whereby increased sclerostin levels cause a decrease in BMD, whereas higher BMD increases sclerostin levels, possibly reflecting a feedback pathway(*15*). Therefore, findings from a sclerostin GWAS are potentially subject to mis-specification of the primary phenotype(*16*)(*17*), with genetic signals being detected which are primarily related to BMD rather than sclerostin.

The goal of the present study was to examine potential safety concerns of sclerostin inhibition on atherosclerosis and its risk factors using an MR approach, based on a set of instruments derived from an updated GWAS meta-analysis of circulating sclerostin, and using 15 atherosclerosis-related diseases and risk factors with available GWAS data, including outcomes such as coronary artery calcification (CAC), type 2 diabetes (T2DM) and lipid/lipoprotein risk factors, which is of interest but understudied. To enable sufficient power to examine causal effects on extra-skeletal phenotypes, we aimed to identify genetic predictors of sclerostin with good instrument strength, incorporating both *cis*- and *trans*-acting variants, having assembled a sample over three times the size of our previous study(*15*). Given *cis*-acting variants are more likely to be specific for the drug target under investigation(*18*), we also aimed to include sensitivity analyses in which MR analyses were restricted to *cis*-acting variants.

## RESULTS

### Summary of study design

**Figure 1** illustrates the design and participants of this study. We aimed to understand the genetic architectural of sclerostin and the causal role of sclerostin inhibition on atherosclerosis related diseases and risk factors. First, we conducted a GWAS meta-analysis and post-GWAS follow-up analyses of circulating sclerostin in 33,961 European individuals, which including nine cohorts: the Avon Longitudinal Study of Parents and Children (ALSPAC), Die Deutsche Diabetes Dialyse Study (4D), The Gothenburg Osteoporosis and Obesity Determinants (GOOD), and the MANOLIS cohort, Fenland(*19*), INTERVAL(*20*), Trøndelag health study (HUNT)(*21*)(*22*)(*23*), the Osteoarthritis Initiative (OAI)(*24*)(*25*)(*26*)(*27*), the Ludwigshafen Risk and Cardiovascular Health (LURIC)(*28*)(*29*). The cohort details were included in **Supplementary Note 1**. Second, we conducted MR analyses of circulating sclerostin using genetic instruments from both *cis* and *trans* regions (**Supplementary Table 1**) as well as from *cis* region only (**Supplementary Table 2**). The outcomes are 15 atherosclerosis related diseases and risk factors (**Supplementary Table 3**). The bi-directional MR was further conducted for the 15 atherosclerosis related diseases and risk factors (**Supplementary Table 4**) on sclerostin.

**Figure 1.**
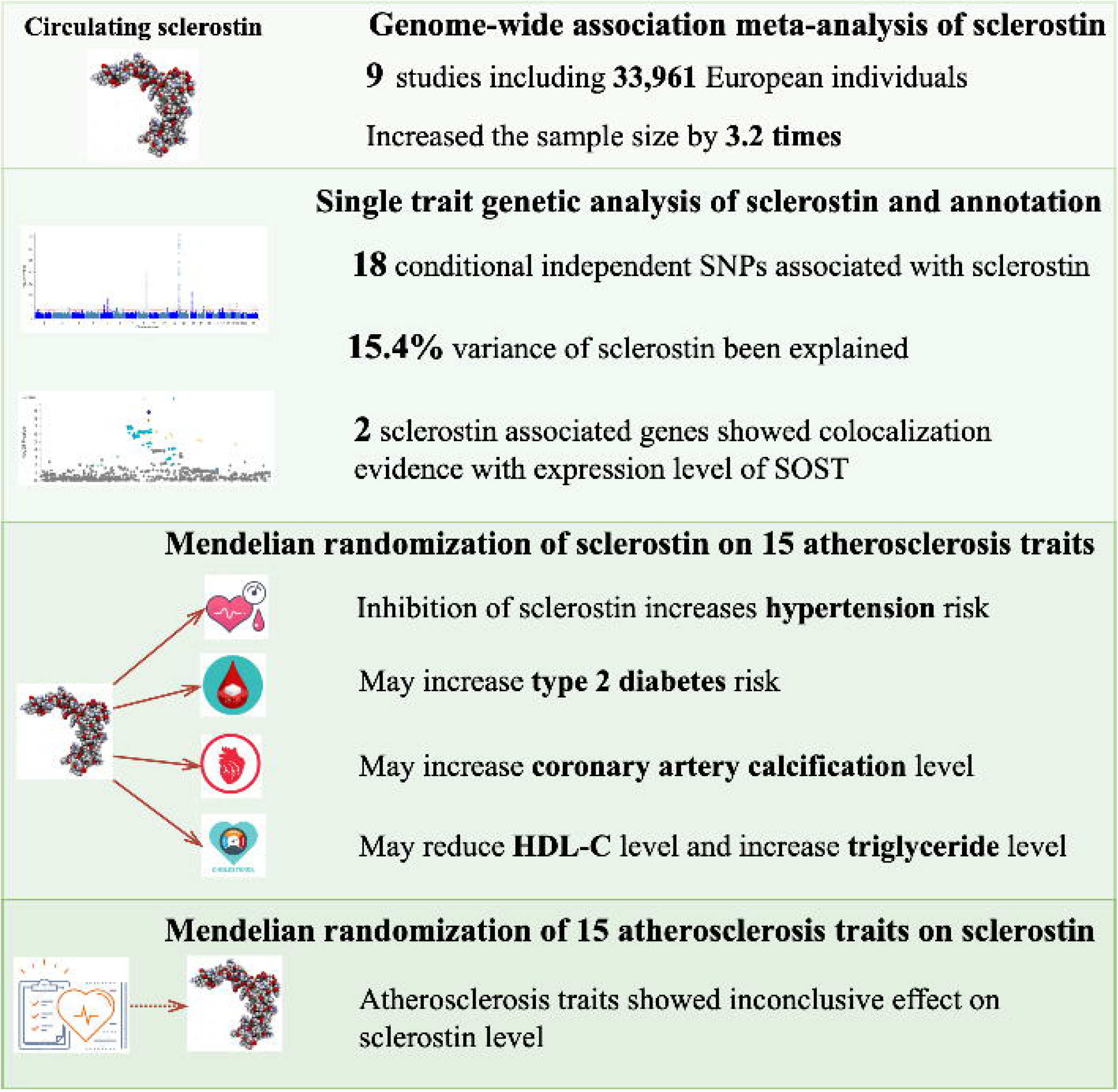
Summary of the design and results of the current study. This study included four major components: (1) meta-analysis of genome-wide association study of circulating sclerostin; (2) signal genetic trait analysis and functional annotation of the top sclersotin signals; (3) Mendelian randomizaiton and genetic correlation analysis of sclerostin on 15 atherosclerosis-related diseases and risk factors traits; (4) bidirectional Mendelian randomization analysis of 15 atherosclerosis-related diseases and risk factors on sclerostin.

### Genome-wide association signals of circulating sclerostin

GWAS results of circulating sclerostin were available in 33,961 European ancestry participants from a meta-analysis of nine cohorts (**Table 1**). **Supplementary Figures 1** and **2** show the Manhattan and QQ plots of association results from the fixed-effects meta-analysis of sclerostin, respectively. There was little evidence of inflation of the test statistics (genomic inflation factor λ=1.082; LD score regression intercept =1.023). Therefore, no genomic control correction was applied to the meta-analysis results. Single trait LD score regression results showed that common variants included in the GWAS meta-analysis explained 15.4% of the phenotypic variance of circulating sclerostin (SNP-based heritability *h^2^*=0.154, SE=0.021, P=3.01×10^-13^).

**Table 1.**
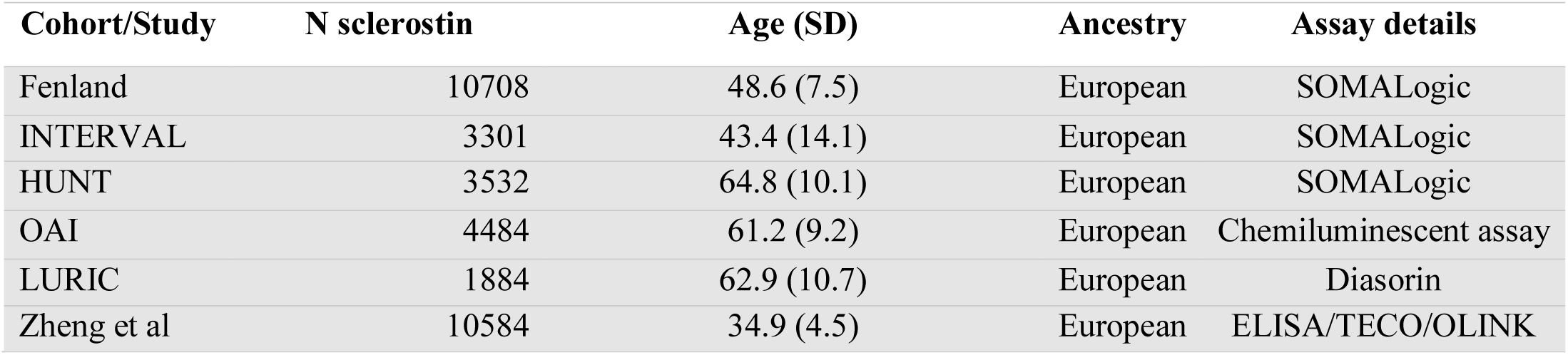
Study information of the cohorts involved in the sclerostin GWAS meta-analysis.

In total, 997 genetic variants were identified to be associated with circulating sclerostin at genome-wide significance. After applying conditional analysis, 18 conditionally independent variants within 15 genomic loci were associated with circulating sclerostin (**Table 2**). The strongest signal, rs215223, was close to the *B4GALNT3* gene (β=-0.136, SD change in circulating sclerostin per A allele, SE=0.008, P=2.44×10^-73^, effect allele frequency=0.405, variance explained=0.89%); the SNP is in perfect LD with the top signal reported in our previous sclerostin GWAS, rs215226(*15*) (**Figure 2A**). One *cis*-acting variant in the *SOST* region, rs66838809, showed a strong association with sclerostin (β=-0.088, SD change in circulating sclerostin per A allele, SE=0.015, P=1.45×10^-9^, effect allele frequency=0.079, variance explained=0.11%; **Figure 2B**). Another variant, rs28929474, in the *SERPINA1* gene region, was associated with circulating sclerostin (β=0.173, SD change in circulating sclerostin per T allele, SE=0.027, P=1.1×10^-10^, effect allele frequency=0.021, variance explained=0.12%; **Figure 2C**). This missense rare variant constitutes the *PiZ* allele, causing alpha-1 anti-trypsin (α1AT) deficiency in homozygous cases(*30*). The variant, rs7143806, in the *RIN3* gene region, was also associated with sclerostin (β=0.053, SD change in circulating sclerostin per A allele, SE=0.010, P=3.35×10^-8^, effect allele frequency=0.181, variance explained by this variant=0.08%; **Figure 2D**). The gene was reported to be associated with lower limb BMD(*31*). The other 12 genomic loci were *FAF1* (rs61781020), *PID1* (rs4973180), *MAP3K1* (rs11960484), *LVRN* (rs34498262 and rs17138656), *SUPT3H* (rs75523462), *LINC00326* (rs34366581), *TNFRSF11B* (rs11995824), *TNFSF11* (rs9594738, rs34136735 and rs665632), *TNFRSF11A* (rs2957124), *JAG1* (rs13042961) and rs6585816 in chromosome 10 (no nearby genes). These include SNPs related to *TNFSF11* and *TNFSF11A*, two well established BMD loci that are assumed to increase sclerostin levels due to greater BMD, and to have no relevance when considering potential therapeutic effects of sclerostin lowering on BMD(*32*).

**Figure 2.**
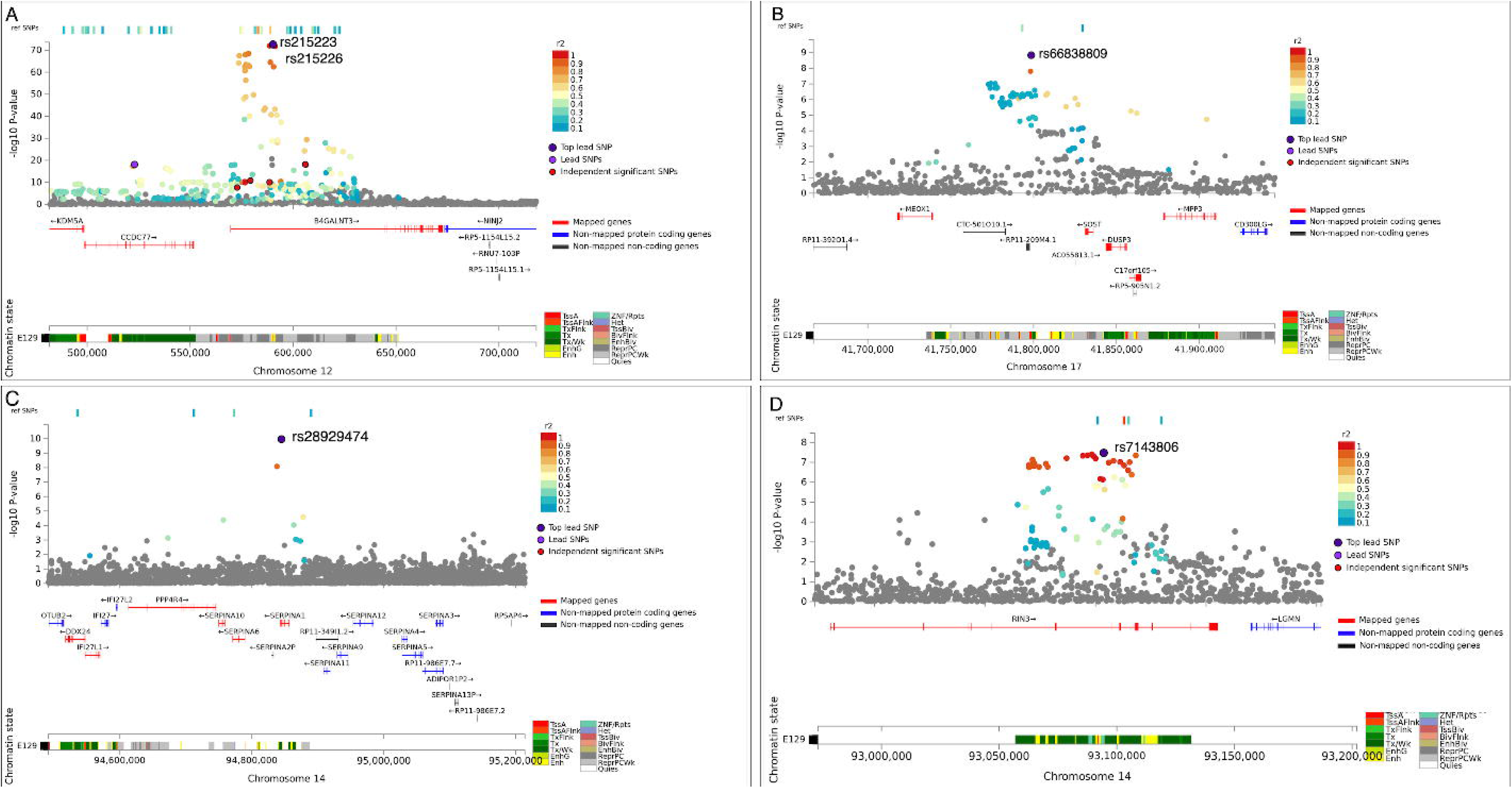
Regional plot for the *B4GLANT3* (A), *SOST* (B), *SERPINA1* (C) and *RIN3* (D) regions. Description of the regulation elements listed in Supplementary Table 13.

**Table 2.** Meta-analysis results for loci that reached genome-wide significance (P < 5 × 10^−8^).

| Locus | SNP | EA | OA | EAF | GENE | Cis/trans | BETA | SE | P | Q | Q P | I2 |
| --- | --- | --- | --- | --- | --- | --- | --- | --- | --- | --- | --- | --- |
| chr1 50566286 | rs61781020 | A | G | 0.049 | *FAF1* | *Trans* | 0.101 | 0.018 | $2.57 \times 10^{-8}$ | 17.825 | 0.058 | 0.439 |
| chr2 229236796 | rs4973180 | T | C | 0.821 | *PID1* | *Trans* | 0.059 | 0.010 | $1.04 \times 10^{-9}$ | 6.691 | 0.754 | 0 |
| chr5 56813327 | rs11960484 | A | G | 0.351 | *MAP3K1* | *Trans* | -0.049 | 0.008 | $1.40 \times 10^{-10}$ | 14.823 | 0.139 | 0.325 |
| chr5 115994797 | rs34498262 | A | G | 0.391 | *LVRN* | *Trans* | 0.065 | 0.008 | $1.41 \times 10^{-17}$ | 10.403 | 0.406 | 0.039 |
| chr5 116013119 | rs17138656 | A | G | 0.124 | *LVRN* | *Trans* | 0.096 | 0.011 | $3.16 \times 10^{-17}$ | 14.273 | 0.161 | 0.299 |
| chr6 45189983 | rs75523462 | T | G | 0.950 | *SUPT3H* | *Trans* | 0.104 | 0.017 | $1.31 \times 10^{-9}$ | 7.424 | 0.685 | 0 |
| chr6 133044782 | rs34366581 | T | G | 0.326 | *LINC00326* | *Trans* | 0.047 | 0.008 | $2.80 \times 10^{-9}$ | 13.204 | 0.212 | 0.243 |
| chr8 119000461 | rs11995824 | C | G | 0.454 | *TNFRSF11B* | *Trans* | 0.100 | 0.007 | $5.62 \times 10^{-41}$ | 16.222 | 0.093 | 0.384 |
| chr10 122342063 | rs6585816 | T | G | 0.209 | / | *Trans* | 0.056 | 0.009 | $7.84 \times 10^{-10}$ | 6.121 | 0.805 | 0 |
| chr12 481093 | rs215223 | A | G | 0.405 | *B4GALNT3* | *Trans* | -0.136 | 0.008 | $2.44 \times 10^{-73}$ | 83.297 | $1.13 \times 10^{-13}$ | 0.880 |
| chr13 42378009 | rs9594738 | T | C | 0.482 | *TNFSF11* | *Trans* | -0.056 | 0.007 | $6.48 \times 10^{-14}$ | 9.217 | 0.512 | 0 |
| chr13 42513606 | rs34136735 | T | C | 0.052 | *TNFSF11* | *Trans* | 0.171 | 0.017 | $5.69 \times 10^{-23}$ | 17.750 | 0.059 | 0.437 |
| chr13 42532378 | rs665632 | T | C | 0.813 | *TNFSF11* | *Trans* | 0.082 | 0.010 | $4.82 \times 10^{-17}$ | 8.502 | 0.484 | 0 |
| chr14 92637384 | rs7143806 | A | G | 0.181 | *RIN3* | *Trans* | 0.053 | 0.010 | $3.35 \times 10^{-8}$ | 13.360 | 0.204 | 0.251 |
| chr14 94378610 | rs28929474 | T | C | 0.021 | *SERPINA1* | *Trans* | 0.173 | 0.027 | $1.10 \times 10^{-10}$ | 5.342 | 0.867 | 0 |
| chr17 43721253 | rs66838809 | A | G | 0.079 | *SOST* | *Cis* | -0.088 | 0.015 | $1.45 \times 10^{-9}$ | 13.369 | 0.147 | 0.327 |
| chr18 62390996 | rs2957124 | A | G | 0.421 | *TNFRSF11A* | *Trans* | -0.057 | 0.008 | $5.97 \times 10^{-14}$ | 11.286 | 0.257 | 0.203 |
| chr20 11231094 | rs13042961 | T | C | 0.955 | *JAG1* | *Trans* | 0.126 | 0.019 | $4.65 \times 10^{-11}$ | 16.398 | 0.037 | 0.512 |

Note: Locus (chromosome and position of the SNP), EA (effect allele), OA (other allele), EAF (effect allele frequency), GENE (nearest gene to the sclerostin associated SNP); Cis/trans (the associated SNP is close to the SOST region [noted as cis] or far away from this region [noted as
trans]); BETA (SD change in serum sclerostin per effect allele), SE (standard error) and P (p-value)). Heterogeneity test (Q (Cochran's Q statistics), Q\_P (Cochran's Q P value), $I^2$ ( $I^2$ statistics))

Results of the random effects meta-analysis were similar to those of the fixed-effect meta-analysis (results not shown). The degree of heterogeneity was low across studies for most of the identified genetic variants (**Table 2**). Conditional analyses on the lead SNP in each association locus yielded one additional independent signal reaching genome-wide significance in the *LVRN* gene region and two additional independent signals in the *TNFSF11* gene region (**Supplementary Table 5**).

### Genetic colocalization analysis of sclerostin association signals with gene expression

For the 18 sclerostin associated variants, we identified four variants [rs215223 (in the *B4GALNT3* region), rs28929474 (in the *SERPINA1* region), rs66838809 (in the *SOST* region) and rs7143806 (in the *RIN3* region)] where sclerostin-increasing alleles were associated with lower eBMD (at P<0.001), whereas in the case of the remaining SNPs, variants were either not associated with eBMD or the sclerostin-increasing alleles were associated with higher eBMD, and were not considered further (**Supplementary Table 5**). We conducted genetic colocalization analysis for rs215223, rs28929474, rs66838809 and rs7143806 to confirm the causal variants were shared between circulating sclerostin levels and gene expression levels of the related genes in tibial artery (data from GTEx v8). The expression of *B4GALNT3* and *SOST* genes showed strong evidence of colocalization with circulating sclerostin levels (colocalization probability=99% and 98%, respectively; **Supplementary Table 6A**). The *SERPINA1* and *RIN3* signal showed weaker evidence of colocalization with sclerostin (colocalization probability=7% and 30%; **Supplementary Figure 3** and **Supplementary Table 6A**). We also confirmed that the *SOST* SNP we identified was associated with altered *SOST* expression in iliac crest bone tissue (**Supplementary Table 6B**)(*33*).

### Bioinformatics functional follow-up

We investigated possible effects of rs215223 (in the *B4GALNT3* region) and rs66838809 (in the *SOST* region) on transcriptional activity. The chromatin accessibility analysis predicted by ChromHMM based on 5 chromatin marks for 127 epigenomes(*34*) identified that the *B4GALNT3* variant (rs215223) overlapped with an enhancer region in osteoblast primary cells (OPCs), and the *SOST* variant (rs66838809) overlapped with an active transcription start site (TSS) in OPCs (see **Figure 2**). The same analysis in other heart and vascular-related tissues showed that the *SOST* variant (rs66838809) also overlapped with an active TSS in three heart tissues. The *SERPINA1* and *RIN3* variants overlapped with weak transcription (**Supplementary Table 7**). Regulatory elements analysis using RegulomeDB(*35*) graded *B4GALNT3*, *SOST* and *RIN3* variants as rank 2B, 3A and 3A, respectively (lower rank implies greater predicted functional impact; **Supplementary Table 5**). The eQTL lookup using STARNET showed that our top hits rs215223 (for) and rs66838809 (for SOST) were associated with gene expression levels of *B4GALNT3* and *SOST* in free internal mammary artery (MAM) respectively (**Supplementary Table 6C**). The gene-set enrichment analysis showed that *RIN3* and *B4GALNT3* were enriched in the same enzyme linked receptor protein signalling pathway (Gene Ontology ID: GO:0007167). SERPINA1 and SOST were enriched in two separate pathways that related to inflammatory response (GO:0072358). In addition, SOST was also enriched in a cardiovascular system development pathway (GO:0072358), where RIN3 was enriched in a pathway that positive regulate immune system process (GO:0002684) (**Supplementary Table 6D**).

### Genetic correlation between sclerostin levels and atherosclerosis-related traits

As expected, genetic correlation analysis between circulating sclerostin using genetic variants across the whole genome revealed a relationship between lower sclerostin and higher eBMD and, to a lesser extent, lower fracture risk (**Supplementary Table S8**). These analyses also showed a genetic overlap of lower sclerostin with increased hypertension risk (r_g_=0.134, P=3.10×10^-3^), but not with any other atherosclerosis-related diseases or risk factors (**Table 3** and **Supplementary Figure 4**).

**Table 3.** Mendelian randomization and genetic correlation analysis results of the effect of sclerostin inhibition on coronary artery calcification, hypertension and type 2 diabetes.

| Exposure | Outcome | Model | N SNPs | Estimate | SE | P | OR | LCI | UCI |
| --- | --- | --- | --- | --- | --- | --- | --- | --- | --- |
| Sclerostin inhibition | Coronary artery calcification | *Cis*-only MR | 5 | 0.740 | 0.210 | $4.27 \times 10^{-4}$** | / | / | / |
|  |  | *Cis+trans* MR | 4 | 0.058 | 0.126 | 0.645 | / | / | / |
| Sclerostin inhibition | Hypertension | *Cis*-only MR | 5 | 0.073 | 0.034 | 0.030 * | 1.080 | 1.010 | 1.150 |
| | | *Cis+trans* MR | 4 | 0.086 | 0.026 | $7.93 \times 10^{-4}$** | 1.090 | 1.037 | 1.146 |
| Sclerostin inhibition | Type 2 diabetes | *Cis*-only MR | 5 | 0.233 | 0.080 | $3.66 \times 10^{-3}$** | 1.262 | 1.079 | 1.477 |
|  |  | *Cis+trans* MR | 3 | 0.034 | 0.138 | 0.804 | 1.035 | 0.789 | 1.358 |

| Trait 1 | Trait 2 | Model | N SNPs | $r_g$ | SE_ $r_g$ | P_ $r_g$ |
| --- | --- | --- | --- | --- | --- | --- |
| Sclerostin inhibition | Coronary artery calcification | Genetic correlation | All SNPs | 0.007 | 0.083 | 0.933 |
| Sclerostin inhibition | Hypertension | Genetic correlation | All SNPs | 0.134 | 0.045 | $3.10 \times 10^{-3}$** |
| Sclerostin inhibition | Type 2 diabetes | Genetic correlation | All SNPs | -0.041 | 0.073 | 0.573 |

Note: Model refers to different statistical methods/models been used. N\_snps means the number of genetic variants been included as predictors for sclerostin. Estimate, SE and P (and $r_g$ , SE\_ $r_g$ , P\_ $r_g$ ) are the association estimates, standard error and P value of the MR (or the genetic correlation analysis). OR, LCI and UCI are the odds ratio and 95% confidence interval of the MR estimates, which is not applicable for the genetic correlation analysis. Importantly, the Mendelian randomization and genetic correlation analyses have different assumptions therefore the effect estimate is not directly comparable. We listed them in the same table to compare the direction of effects and the P value estimates across the two approaches.
\* Association reached marginal significance threshold of $\alpha=0.05$
\*\* Association reached Bonferroni-corrected significance threshold of $\alpha=4.17 \times 10^{-3}$

### Sclerostin instruments and effects of lower sclerostin on risk of atherosclerosis-related diseases and risk factors

MR analyses of the effect of lower sclerostin levels on atherosclerosis risk combined the *SOST cis* variants with the *B4GALNT3, SERPINA1* and *RIN3 trans* variants identified above. For all four SNPs, the alleles associated with lower circulating sclerostin levels were associated with increased eBMD and reduced fracture risk **(Figure 3A)**. Compared to the other three variants, the *SOST* SNP showed a disproportionately strong association with eBMD (**Supplementary Table 1A**), relative to its association with circulating sclerostin (**Figure 3B**). Together, these four SNPs explained 1.21% of the variance in circulating sclerostin and provided a strong genetic instrument (F-statistic 89.8, an F statistic of at least 10 is indicative of evidence against weak instrument bias) **(Supplementary Table 1).** For the remaining 14 sclerostin variants, five variants were not associated with eBMD, where the other nine variants showed directionally similar effects on eBMD and sclerostin. These variants did not fit with our selection criteria and were therefore excluded from the instrument list (**Supplementary Figure 5**).

**Figure 3.**
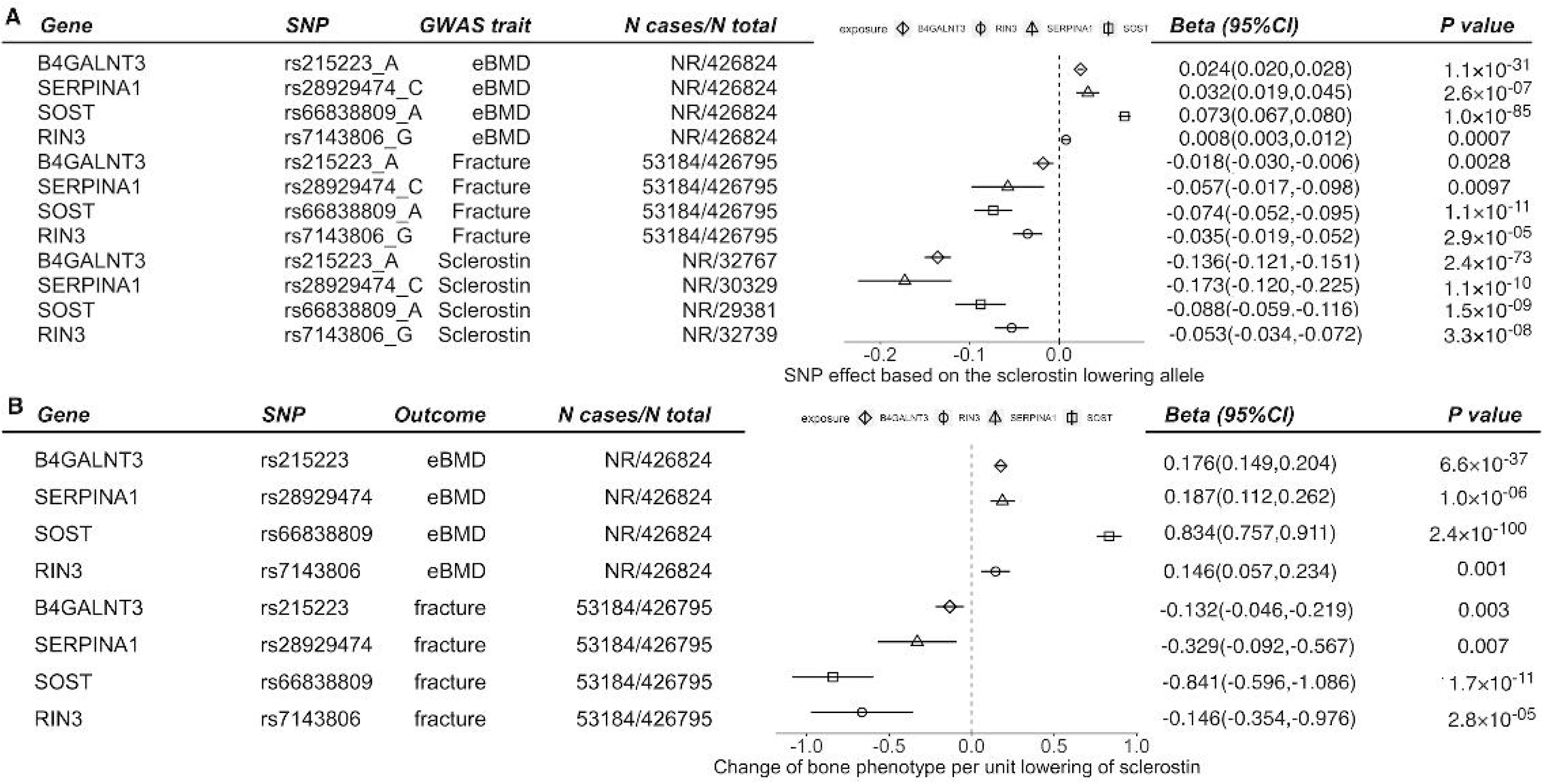
Genetic effect of sclerostin-associated SNPs on eBMD and fracture. (A) Genetic effects of four variants on sclerostin, fracture, and eBMD. The alleles presented in the plot are the sclerostin-lowering alleles. Different colour refers to the three traits been plotted. (B) Genetic effect of three sclerostin variants on eBMD and fracture scaled to standard deviation unit of sclerostin reduction.

Using these four conditionally independent SNPs to evaluate causal effects of lower sclerostin levels on atherosclerosis-related diseases and risk factors (Bonferroni-corrected threshold=5.15×10^-3^), a predicted lower circulating sclerostin was found to be associated with an increased risk of hypertension (OR per SD decrease in sclerostin= 1.09, 95% CI=1.04 to 1.15, P=7.93×10^-4^) **(Figure 4A).** Sensitivity analyses including MR-Egger and a heterogeneity test suggested little evidence of horizontal pleiotropy (Egger regression intercept=-0.003, P=0.27) or heterogeneity (Cochran’s Q=2.85, P=0.42; **Supplementary Table 9A**). In contrast, little evidence for a causal effect of lower sclerostin on any other atherosclerosis-related disease or risk factor was identified. Given the low power to detect pleiotropy using the above methods due to the limited number of genetic instruments available, we conducted a proteome-wide association scan of the four genetic variants to further examine potential pleiotropy. All variants, except rs28929474 within the *SERPINA1* gene region, showed little evidence of association with any other proteins, where rs28929474 was associated with additional 27 proteins (**Supplementary Table 9B**).

**Figure 4.**
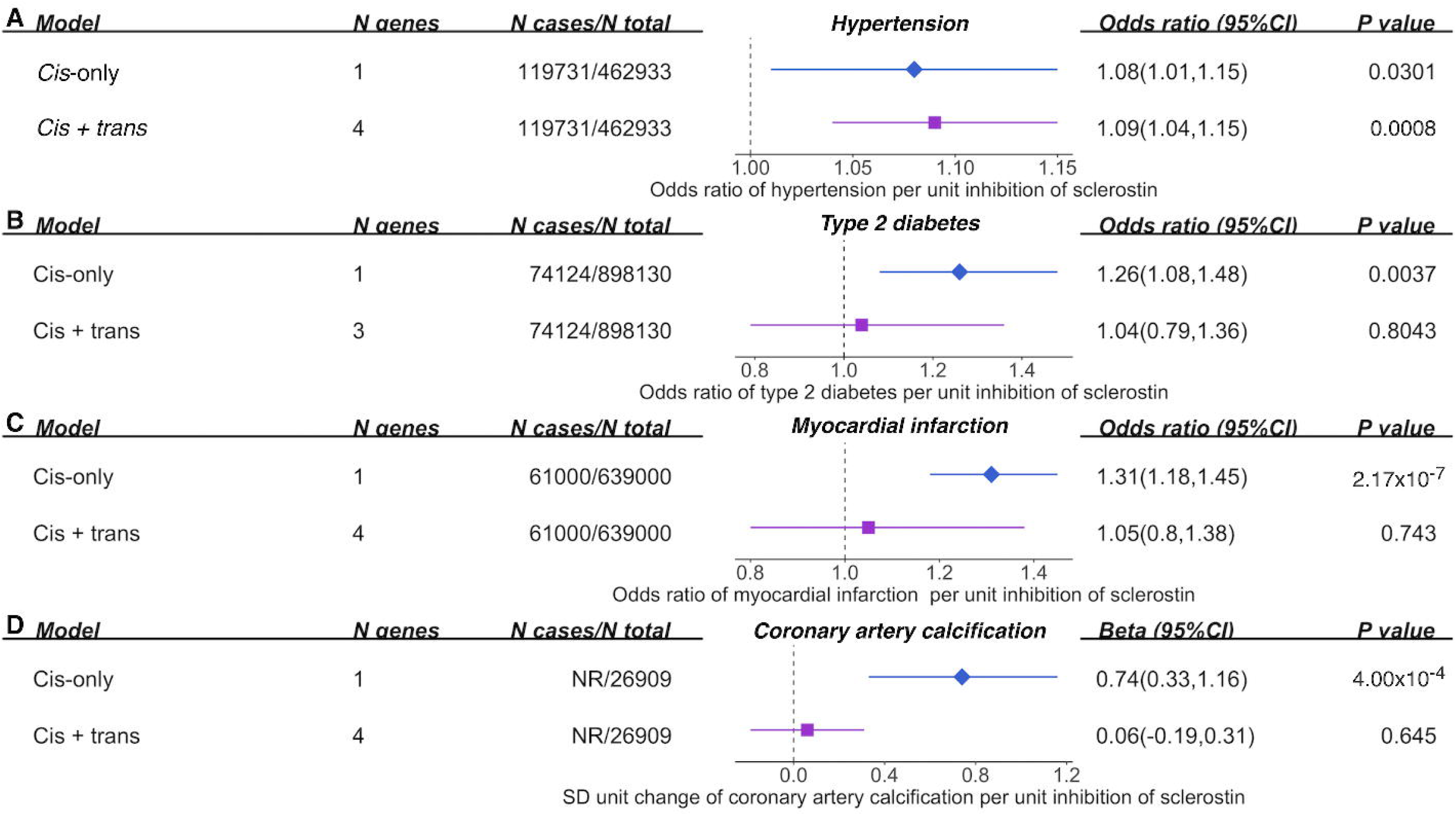
Causal effects of circulating sclerostin inhibition on hypertension, type 2 diabetes and coronary artery calcification using cis-only and cis+trans genetic predictors of sclerostin. (A) causal effect of sclerostin inhibition on hypertension risk; (B) causal effect of sclerostin inhibition on type 2 diabetes risk; (C) causal effect of sclerostin inhibition on coronary artery calcification.

In further analyses using *cis*-only instruments, we observed that the five correlated variants (rs66838809, rs1107747, rs4793023, rs80107551, rs76449013) together explained 0.4% of the variance in circulating sclerostin and had acceptable instrument strength (F-statistic 24.8) (**Supplementary Table 2**). The *cis*-only analysis identified potential adverse effects of sclerostin inhibition on risk of hypertension (OR=1.08, 95% CI=1.01 to 1.15, P=0.03; **Figure 4A**), T2DM (OR=1.26, 95% CI=1.08 to 1.48, P=0.004; **Figure 4B**) and MI (OR=1.31, 95% CI=1.183 to 1.45, P=2.17×10^-7^; **Figure 4C**). Genetically predicted lower sclerostin was associated with higher levels of CAC (β=0.74; 95% CI=0.33 to 1.15; P=4.27×10^-4^; **Figure 4D**). In addition, lower sclerostin in the *cis*-only analyses showed potential harmful effect on AAC and CAD, but with very wide confidence intervals (**Supplementary Table 10A**). In addition, sclerostin showed effects on four of the five lipids and/or lipoproteins in the *cis*-only analysis but with little evidence of MR effects in the combined *cis* and *trans* analysis (**Figure 5A** and **Supplementary Table 10A**). This could be partly caused by the LD between the *cis SOST* variant (rs4793023) and the top-associated variant with mRNA *CD300LG* expression (rs72836567; LD r^2^=0.22 in the 1000 Genomes EUR population)(*36*), where *CD300LG* is known to be strongly associated with lipid measures(*37*). As a sensitivity analysis, we excluded rs4793023 from the genetic predictor list and ran the *cis*-only MR using the remaining four predictors. The results suggested that decreased sclerostin levels reduced HDL-C levels and increased triglycerides levels, whereas the MR effects on other lipids/lipoproteins were attenuated after this adjustment (**Supplementary Table 10B**). Heterogeneity analysis of MR estimates of each genetic instrument suggested little evidence of heterogeneity across the five genetic instruments (Cochran’s Q test P>0.05 for these four lipids and/or lipoproteins measures; **Supplementary Table 10A** and **10B**).

**Figure 5.**
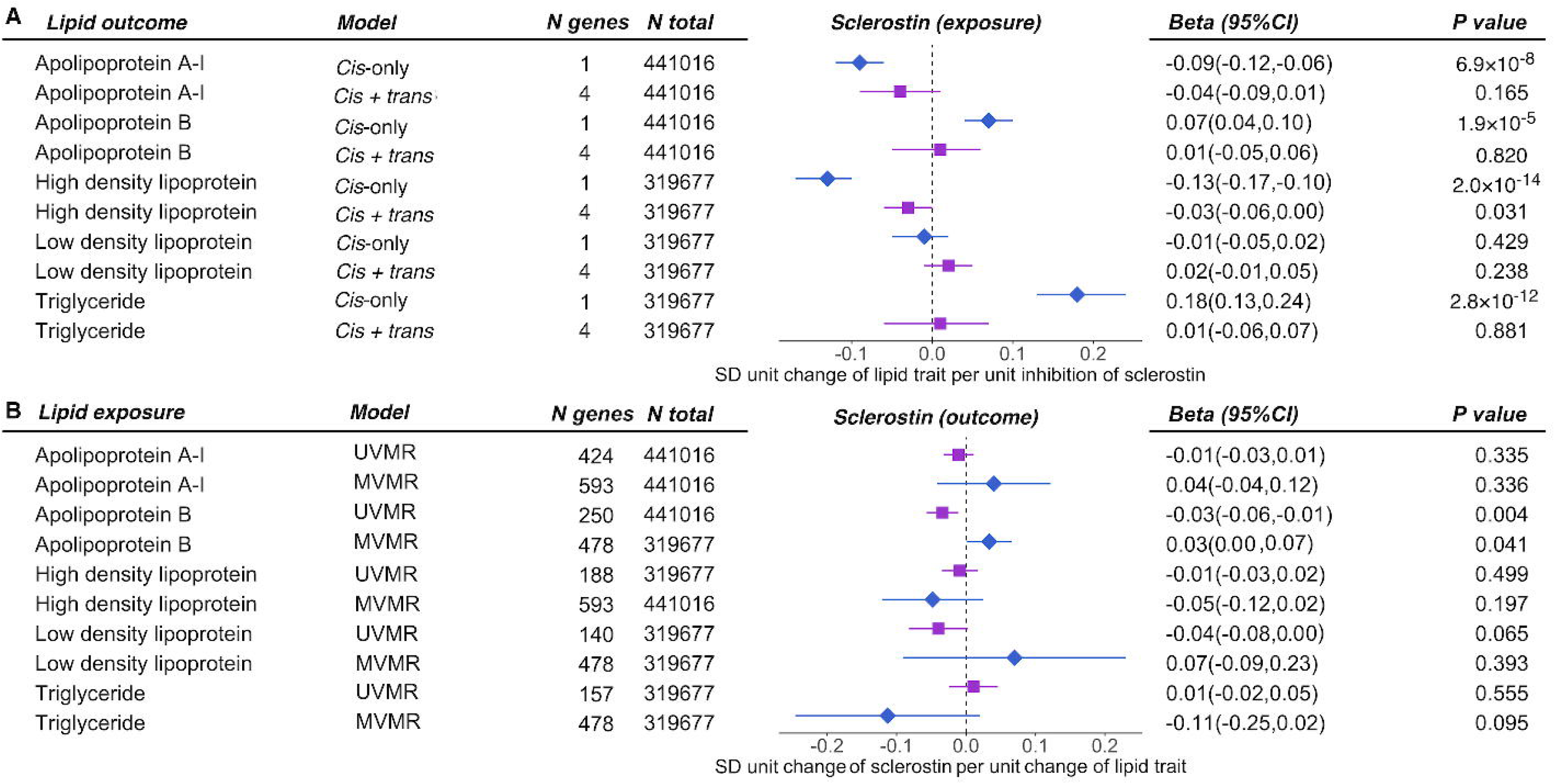
Bidirectional causal effects between circulating sclerostin and five lipids traits. (A) causal effect of sclerostin inhibition on five lipid traits using cis-only and cis+trans instruments; (B) causal effect of five lipid traits on sclerostin using univariate (UVMR) and multivariable Mendelian randomization (MVMR).

### Effects of atherosclerosis-related diseases and risk factors on circulating sclerostin

We further conducted bidirectional MR(*38*) to evaluate the potential reverse causality of atherosclerosis-related diseases and risk factors on circulating sclerostin. We used the 15 atherosclerosis-related diseases and risk factors as exposures, of which nine had valid predictors to conduct bidirectional MR (**Supplementary Table 4**; small vessel disease had no valid genetic predictors and was therefore excluded from this analysis). Circulating sclerostin was treated as the outcome. IVW results using 226 T2DM-associated variants showed a marginal positive relationship for liability of T2DM on sclerostin (β=0.02, SD change in sclerostin per unit increase of risk score of T2DM, 95%CI= 0.001 to 0.045, P=0.04; **Supplementary Table 11A**). The IVW results showed that apoB was negatively associated with sclerostin levels (β=-0.03, 95%CI=-0.01 to -0.06, P=3.67×10^-3^). However, the multivariable MR including apoB, LDL-C and triglycerides in the same model suggested that increased apoB levels increased sclerostin levels (β=0.03, 95%CI=0.001 to 0.07, P=0.041; **Figure 5B** and **Supplementary Table 11B**). No other atherosclerosis-related disease or risk factor showed a reverse effect on sclerostin (**Supplementary Table 11A**). The MR-Egger intercept test did not suggest evidence of directional pleiotropy. The heterogeneity test showed weak evidence for any heterogeneity of the causal estimates (**Supplementary Table 11A**). The Steiger filtering analysis further confirmed that the sclerostin instruments were likely to first change the sclerostin level and then influence the atherosclerosis outcomes as a causal consequence (**Supplementary Table 11C**).

## DISCUSSION

We have presented findings from an updated GWAS meta-analysis of circulating sclerostin, with three times the sample size of our previous study(*15*). We identified 18 sclerostin-associated variants, of which four in the *SOST*, *B4GALNT3, RIN3* and *SERPINA1* genes provided useful genetic instruments for determining the causal effects of lower sclerostin levels on atherosclerosis-related diseases and risk factors based on inverse relationships between sclerostin levels and BMD. The *B4GALNT3* result replicated the top hit, rs215226, from our previous study; *RIN3* and *SERPINA1* are novel sclerostin-associated *trans* signals, and the *SOST* finding represents a strong *cis* signal which was only marginal in our previous study. MR analyses using these four SNPs as a combined *cis* and *trans* genetic instrument suggested that lower sclerostin levels increase the risk of hypertension, however there was little evidence of a relationship with other atherosclerosis-related diseases or risk factors. On the other hand, sensitivity analyses using a *cis*-only genetic instrument suggested that lower sclerostin levels increase the risk of hypertension, T2DM and MI and increase the extent of CAC. In addition, *cis*-only analyses suggested that lower sclerostin levels reduce HDL-C and apoA-I, and increase apoB and TG; effects on HDL-C and TG persisted following exclusion of the *SOST* variant rs4793023 (in LD with rs72836567 in the adjacent *CD300LG* gene previously found to be associated with lipid measures)(*37*).

Lower sclerostin levels had similar predicted effects on hypertension risk using both *cis*-only and combined *cis* and *trans* instruments, and we observed evidence of genetic correlation between sclerostin inhibition and hypertension using variants across the whole genome, without evidence of reverse causality. Together, these findings provide reasonable evidence of a causal effect of sclerostin on risk of hypertension. *cis*-only analyses suggested additional causal effects of lower sclerostin levels on atherosclerosis-related diseases and risk factors, and in particular that inhibition of sclerostin level increases risk of MI as a consequence of greater CAC. That said, relationships between CAC and clinical events are potentially complex, with some evidence suggesting that CAC reflects the presence of stable atherosclerotic plaques with a reduced risk of coronary artery occlusion compared with uncalcified plaques(*39*)(*40*)(*41*). Bidirectional analyses broadly supported a causal effect of sclerostin inhibition on increased risk of atherosclerosis-related diseases and risk factors, as opposed to vice versa. That said, in addition to a causal effect of lower sclerostin levels on risk of T2DM in *cis*-only analyses, there was marginal evidence for a causal effect of T2DM on sclerostin. Moreover, whereas the *cis* instrument suggested a causal effect of sclerostin inhibition on CAC and MI, there was no equivalent effect on AAC.

Cis instruments are more likely to directly link with biology, which aligns with our finding that *cis*-only analyses identified more extra-skeletal effects of sclerostin. *Trans* instruments are, by their nature, more likely to be pleiotropic, and as a result, they may include additional pathways responsible for this apparent divergence in effects on eBMD and extra-skeletal pathways. Additionally, *cis* variants may be more predictive of tissue sclerostin levels responsible for mediating biological effects. Based on eQTL data using bone tissue, the *cis* signal is predicted to alter expression and hence local levels of sclerostin in bone. Osteocytes, embedded within bone, are the primary source of sclerostin, which then circulates locally through canaliculi to modulate the activity of other bone cells, including osteoblasts, leading to changes in bone mass and strength(*42*). Accordingly, the *cis* signal is expected to alter circulating levels of sclerostin through exchange between bone tissue and the circulation. In contrast, we previously hypothesised that the *trans* signal, *B4GALNT3*, replicated in the present study, primarily influences circulating sclerostin levels by affecting plasma clearance due to altered protein glycosylation(*15*). Hence, any changes in tissue sclerostin levels resulting from the *B4GALNT3 trans* signal are likely secondary to altered circulating levels, rather than local production. Therefore, by its nature, the B4GALNT3 *trans* signal is expected to produce smaller changes in tissue sclerostin levels compared to a *cis SOST* signal, leading to a weaker effect on eBMD.

That the *SOST cis* signal is likely to produce greater increases in tissue sclerostin levels compared to *trans* signals may also explain why the *cis*-only analyses predicted more extra-skeletal effects of sclerostin lowering compared to the *cis+trans* analyses. Sclerostin is also expressed in vascular tissues including at sites of vascular calcification(*43*)(*44*), suggesting any effects of sclerostin on vascular tissues may also involve local sclerostin expression. Such an effect is likely mediated by sclerostin’s well recognised action as a WNT inhibitor(*45*), given the contribution of WNT signalling to the development of atherosclerosis(*46*).

Pharmacokinetic studies suggest that romosozumab is largely retained within the circulation(*13*), in-keeping with the relatively large size of a monoclonal antibody. That said, the pharmacological action of romosozumab, involving neutralisation of sclerostin activity in bone tissue, depends on the antibody penetrating skeletal tissue after systemic administration, which is likely to involve convection or endocytosis/pinocytosis via endothelial cells(*47*). To the extent that effects of romosozumab on CVD risk also involve local tissue penetration, a *cis* instrument reflecting tissue levels of sclerostin may be more likely to predict effects of romosozumab on CVD risk than a *trans* instrument more closely linked to systemic levels.

There have also been several previous observational studies examining associations between circulating sclerostin and atherosclerosis related diseases and risk factors, of which the largest was our recent study of over 5000 participants across two cohorts(*7*). We found that decreased sclerostin levels increased risk of T2DM and triglyceride levels and reduced HDL-C levels; higher sclerostin was also associated with an increased severity of CAD as measured on angiogram, and an increased risk of death from cardiac disease during subsequent follow up(*7*). These observed associations suggest causal effects in the opposite direction to those predicted by our MR analyses, particularly in analyses restricted to the *cis* instrument. Interestingly, directionally opposite effects have also been observed in the case of eBMD and atherosclerosis risk, with a protective effect found in an observational analysis but a harmful effect predicted by MR analyses(*48*). The latter finding also raises the possibility that any effect of sclerostin inhibition on atherosclerosis risk might be an indirect consequence of increased BMD, as opposed to a specific effect of sclerostin. However, arguing against this suggestion, there is little evidence that other therapeutic agents for osteoporosis acting to increase BMD affect atherosclerosis risk, apart from strontium ranelate for which the European Medicines Agency issued a warning, restricting use in those with a high risk of CVD(*49*).

Two previous studies have used MR approaches to examine causal effects of sclerostin inhibition on atherosclerosis and related risk factors. Bovijn *et al.* reported that two conditionally independent *SOST* SNPs, selected on the basis of their association with eBMD in a previous UK Biobank GWAS (of which one SNP also showed evidence of colocalization with lower *SOST* mRNA expression in tibial artery tissue), predicted higher risk of MI and/or coronary revascularization, major cardiovascular events, hypertension, and T2DM(*12*). Our findings, using the *cis*-only instrument for circulating sclerostin, are consistent with these observations. In contrast, Holdsworth *et al.* found no association between gene expression level of *SOST* in tibial artery/heart tissue and CVD risk, using three cis SOST eQTLs as instruments(*14*). Though one *SOST* SNP (rs9899889, in low LD with the *SOST* SNPs used here (see **Supplementary Table 12**) was associated with lower triglyceride levels and higher HDL cholesterol and apoA levels, in contrast to our findings, this signal was entirely explained by LD with the adjacent *CD300LG* gene known to be associated with lipid measures(*37*). Despite the distinct methods used to proxy sclerostin inhibition, our *cis* instrument was in strong LD with those used in these other studies. Indeed, our *cis* instrument shared an identical SNP with the Holdsworth study (see **Supplementary Table 12**).

In terms of other *trans*-acting pathways, we identified a further glycosylation enzyme, *GALNT1*, as being associated with circulating sclerostin levels in our earlier GWAS. However, this association did not replicate in the present expanded GWAS. On the other hand, we identified two new *trans* signals for sclerostin, *RIN3* and *SERPINA1*. Previous GWASs have identified *RIN3* in association with lower limb and total BMD in children(*31*), and Paget’s disease of bone(*50*). Though altered sclerostin levels could conceivably underlie this association with childhood BMD, this is less likely to apply to Paget’s disease, which is primarily an osteoclast disorder. Homozygosity of *SERPINA1* underlies deficiency of α1AT, a glycoprotein mostly produced by the liver, which serves to protect lung tissue from tissue damage caused by proteases released from neutrophils. The loss of function allele was associated with higher sclerostin levels, and the mechanisms underlying this genetic association are unclear. α1AT deficiency causes early-onset chronic obstructive pulmonary disease (COPD)(*51*), and whereas rs28929474 heterozygosity has been associated with increased human height(*51*), we are not aware of any previous findings relating α1AT to BMD or risk of osteoporosis. Given the lack of evidence of colocalization, it is also possible that a different gene was responsible for the genetic signal identified at this locus.

In terms of strengths, the present study had sufficient sample size to clearly detect a *cis (SOST)* signal, and our genetic instrument successfully accounted for bidirectional effects between sclerostin and BMD, by removing *trans* SNPs with the same direction of effect on sclerostin and eBMD. Our MR of sclerostin effects on atherosclerosis-related diseases and risk factors used circulating protein level of sclerostin as the exposure, which may predict adverse effects from sclerostin antibody inhibition more accurately than previous studies using BMD or *SOST* arterial expression as exposures. Finally, since genetic predictors in the *cis*- and/or *trans*-acting regions may yield different causal estimates on outcomes, we considered these separately. In terms of weaknesses, though postmenopausal women are the main target group for osteoporosis treatments such as romososumab, we were only able to examine predicted effects of sclerostin inhibition in males and females combined, due to the lack of availability of sex-specific sclerostin GWAS dataset. In addition, the different cohorts used distinct methods to measure sclerostin, with the over half providing sclerostin measures through the SomaLogic platform, while the other half used a specific ELISA. However, despite these methodological differences, there was little evidence of heterogeneity of genetic associations between cohorts.

In conclusion, our updated GWAS meta-analysis of circulating sclerostin now identified a robust *cis* (*SOST*) signal, replicated our previous *B4GALNT3* signal, and identified new *trans* signals in the *RIN3* and *SERPINA1* genes. To predict adverse effects of sclerostin inhibition, these signals were combined to provide a *cis+trans* instrument for an MR analysis of effects of lower sclerostin levels on atherosclerosis-related diseases and risk factors. Genetically predicted lower sclerostin levels were found to increase the risk of hypertension, a relationship that was supported by the finding of an inverse genetic correlation between sclerostin and hypertension at the genome-wide level, and the lack of any evidence of reverse causality. Analyses based on the *cis* (*SOST*) instrument alone found a similar causal effect of lower sclerostin levels on hypertension risk, and additionally suggested causal effects on risk of MI and T2DM, increased CAC, reduced HDL-C and increased apoB and TG levels. To the extent that genetically predicted lower lifelong exposure to sclerostin shares consequences with pharmacological inhibition over 12 months, our results underscore the requirement for strategies to mitigate potential adverse effects of romosozumab treatment on atherosclerosis and its related risk factors.

## Materials and Methods

### GWAS Meta-analysis of sclerostin

Sclerostin measures in the nine cohorts were standardized to SD units. Each cohort ran a GWAS across all imputed or sequenced variants. Age and sex and the first 10 principal components (PCs) were included as covariates in all models (except INTERVANL and LURIC). For the INTERVAL study, age, sex, duration between blood draw and processing (binary, ≤1 day / >1 day) and the first three PCs were included in the genetic association model. The Fenland, ALSPAC, 4D, and GOOD cohorts were imputed using the Haplotype Reference Consortium (HRC) V1.0 reference panel (MANOLIS employed whole-genome sequencing). Linear mixed models BOLT-LMM and GEMMA were applied to the ALSPAC and MONOLIS cohorts, respectively, to adjust for cryptic population structure and relatedness. Linear regression was performed using BGENIE (v1.3)(*52*) in the Fenland study. For the INTERVAL study, a linear regression model was applied using genotype data imputed by a combined 1000 Genomes Phase 3-UK10K reference panel. The LURIC cohort was imputed to the 1000 Genomes Phase 1 reference panel with linear regression performed in PLINK v1.90(*53*) using eight ancestry components as covariates to control for population substructure. In HUNT, the GWAS for protein was performed using rank transformed residuals of protein adjusted for age, sex, batch effect, phase effect and principal components from 1-20.

We standardized the genomic coordinates to be reported on the NCBI build 37 (hg19), and alleles on the forward strand. Summary level quality control was conducted for Europeans only in EasyQC(*54*). Meta-analysis (using a fixed-effect model implemented in METAL(*55*)) was restricted to variants with a minimal sample size >10,000 individuals, MAF >1%, and high imputation quality score (R^2^ >0.8 for variants imputed in MaCH(*56*) and INFO >0.8 for variants imputed in IMPUTE(*57*) (n=11,680,861 variants). Meta-analysed P value lower than 5×10^−8^ was used as a threshold to define genome-wide significant associations. A random effects model meta-analysis was also conducted using GWAMA version 2.2.2(*58*). Heterogeneity was assessed using the I^2^ statistic and Cochran’s Q test.

#### Conditional analysis and genetic fine mapping

We carried out an approximate conditional and joint genome-wide association analysis (GCTA-COJO) to detect multiple independent association signals at each of the sclerostin locus(*59*). SNPs with high collinearity (Correlation r^2^ > 0.9) were ignored, and those situated more than 10 Mb away were assumed to be in complete linkage equilibrium (LD). A reference sample of 8,890 unrelated individuals of ALSPAC mothers was used to model patterns of LD between variants. The reference genotyping data set consisted of the same 11.6 million variants assessed in our GWAS. Conditionally independent variants with P<5×10^-8^ were annotated to the physically closest gene with the hg19 gene range list available in dbSNP (https://www.ncbi.nlm.nih.gov/SNP/).

### Functional mapping and annotation of sclerostin genetic association signals

#### Genetic colocalization of gene expression quantitative trait loci (eQTLs) and the sclerostin signals

We investigated whether the SNPs influencing serum sclerostin level were driven by *cis*-acting effects on transcription by evaluating the overlap between the sclerostin-associated SNPs and eQTLs within 500kb of the gene identified, using data derived from all tissue types from GTEx v8(*36*). Evidence of eQTL association was defined as P < 1×10^-4^ and evidence of overlap of signal was defined as high LD (r^2^ ≥ 0.8) between eQTLs and sclerostin-associated SNPs in the region. Where eQTLs overlapped with sclerostin-associated SNPs, we used genetic colocalization analysis(*60*) to estimate the posterior probability (PP) of each genomic locus containing a single variant affecting both circulating sclerostin and gene expression levels in different tissues.

We used Functional Mapping and Annotation of Genome-Wide Association Studies (FUMA), an integrative web-based platform (http://fuma.ctglab.nl), containing information from 18 biological data repositories and tools, to characterise the genetic association signals of sclerostin. According to: (i) functional consequences on gene functions, (ii) mapped genes and biological pathways, and (iii) associations with other phenotypes. The FUMA pipeline has been described in detail elsewhere(*61*). First, we applied the basic plotting function of FUMA to create Manhattan and QQ plots of our sclerostin GWAS meta-analysis results as well as regional plots for top loci. We then applied FUMA’s SNP2GENE function, which used the conditionally independent significant SNPs, and annotated the functional consequences of these variants on gene functions (i.e., altering expression of a gene, affecting a binding site or changing the protein structure). Functionally annotated SNPs were subsequently mapped to genes based on functional consequences by (i) physical position on the genome (positional mapping), (ii) eQTL associations (eQTL mapping), and (iii) 3D chromatin interactions (chromatin interaction mapping). Gene-based/gene-set analyses using MAGMA were carried out to summarize SNP associations at the gene level and associate the set of genes to biological pathways. For each sclerostin-associated locus, we identified all SNPs in high LD with the top signal (LD r^2^>0.8) and characterized their DNA features and regulatory elements in non-coding regions of the human genome using RegulomeDB (http://www.regulomedb.org/)(35), as implemented in FUMA. In addition, we estimated the chromatin accessibility of genomic regions (every 200bp) for the top loci (*SOST* and *B4GLANT3*) using 15 categorical states predicted by ChromHMM (*34*) based on five chromatin marks for 127 epigenomes (lower state with higher accessibility).

For the top genes been identified as associated with circulating sclerostin and eBMD, we searched their eQTLs in 7 tissues related to cardio-metabolic phenotypes (which including blood, free internal mammary artery [MAM], atherosclerotic aortic root [AOR], subcutaneous fat [SF], visceral abdominal fat [VAF], skeletal muscle [SKLM], and liver [LIV]) as well as gene-set enrichment using the STARNET web app(*62*).

### LD score regression analyses

#### Estimation of SNP heritability using LD score regression

To estimate the amount of genomic inflation in the data due to residual population stratification, cryptic relatedness, and other latent sources of bias, we used LD score regression(*63*). LD scores were calculated for all high-quality SNPs (i.e., INFO score [or R^2^] > 0.9 and MAF > 0.1%) from the meta-analysis. We further quantified the overall SNP-based heritability with LD score regression using a subset of 1.2 million HapMap SNPs (SNPs in the major histocompatibility complex [MHC] region were removed due to complex LD structure).

#### Estimation of genetic correlations using LD Hub

To estimate the genetic correlation between reduced sclerostin level and 12 atherosclerosis-related diseases and risk factors and two bone phenotypes, we used a platform based on LD score regression as implemented in the online web utility LD Hub(*64*). This method uses the cross-products of summary test statistics from two GWASs and regresses them against a measure of how much variation each SNP tags (i.e., its LD score). Variants with high LD scores are more likely to contain more true signals and thus provide a greater chance of overlap with genuine signals between GWASs. The LD score regression method uses summary statistics from the GWAS meta-analysis of sclerostin and the atherosclerosis-related diseases and risk factors/bone phenotypes, calculates the cross-product of test statistics at each SNP, and then regresses the cross-product on the LD score. The small vessel disease data had the heritability estimate out-of-bounds and was therefore removed.

### Mendelian randomization

#### Selection of genetic predictors for sclerostin

From the 18 conditionally independent sclerostin variants identified (**Supplementary Table 1A**), we selected valid genetic predictors of sclerostin for the MR using three further criteria: (i) the genetic variants showed predicted effects of sclerostin on BMD estimated using ultrasound in heel (eBMD, data from UK Biobank; single SNP MR P value of sclerostin on eBMD<0.001); (ii) the sclerostin reducing alleles of the genetic variants were associated with increased BMD level (i.e., these variants showed a negative Wald ratio(*65*) for sclerostin on BMD). The final set of four genetic variants after applying these two additional criteria are listed in **Supplementary Table 1B**. The analysis using these four variants is noted as the *cis* and *trans* analysis.

Due to the greater relevance of the *cis*-acting variants, we conducted a sensitivity analysis using genetic variants restricted to *cis*-acting variants (defined as ±500 kb genomic region from the leading *SOST* SNP) (noted as the *<u>cis</u>*<u>-only analysis)</u>. Of the 41 SNPs associated with circulating sclerostin (at a regional-wide association threshold<1×10^-6^) in the *SOST* region (±500 kb genomic region from rs66838809), LD clumping identified five correlated SNPs with LD r^2^<0.8 (**Supplementary Table 2**). Such an LD r^2^ threshold was used here to avoid multi-collinearity caused by SNPs in very high LD. These correlated instruments were used in a generalised MR model that considered LD among instruments (more details in later section). Instrument strength was evaluated using F-statistics.

#### Outcome selection

For the MR analysis estimating the potential adverse effects of sclerostin inhibition, we selected eight atherosclerosis-related diseases and seven atherosclerosis-related risk factors as primary outcomes (**Supplementary Table 3**). This list comprised two endpoints related to ischaemic heart disease (coronary artery disease (CAD) and MI), four stroke endpoints (ischemic stroke, cardioembolic stroke, large vessel disease, small vessel disease), two measures of arterial calcification [CAC, abdominal aortic calcification (AAC)], hypertension, T2DM, and five lipids/lipoproteins risk factors [low density lipoprotein (LDL-C), high density lipoprotein (HDL-C), triglycerides, apolipoprotein A-I (apoA-I), and apolipoprotein B (apoB)]. After applying PhenoSpD(*66*), which takes into account the correlation between the 15 atherosclerosis-related diseases and risk factors, the number of independent tests was 9.7 (Bonferroni corrected threshold=5.15×10^-3^). We looked up GWAS results in datasets non-overlapping to those used for the sclerostin GWAS, namely the T2DM GWAS from Mahajan *et al.* (N cases=74,124, N controls=824,006)(*67*); four stroke GWASs from the METASTROKE consortium (N cases=10,307, N controls=19,326)(*68*), the CAD GWAS from Van der Harst *et al.* (N cases=34,541, N controls=261,984)(*69*), the MI GWAS from Hartiala *et al.* (N cases=61000, N controls=578000)(*70*), the CAC GWAS from Kavousi *et al.* (N=26,909 CHARGE participants with CAC score)(*71*); the hypertension, lipids and lipoproteins GWASs from UK Biobank IEU GWAS data release (N hypertension cases=119,731, N controls=343,202; N lipids/lipoproteins=441,016)(*72*) and the AAC GWAS from Malhotra *et al.* (sample size=9,417)(*73*).

#### Mendelian randomization of sclerostin on atherosclerosis-related phenotypes

For the *cis* and *trans* analysis, we applied a set of two-sample MR approaches [inverse variance weighted (IVW), MR-Egger, weighted median, single mode estimator and weighted mode estimator](*74*) to estimate the effect of circulating sclerostin on the 15 atherosclerosis-related diseases and risk factors. Although we had a small number of relevant variants available for this analysis, we still used the MR-Egger intercept term as an indicator of potential directional pleiotropy(*75*). Heterogeneity analysis of the instruments was conducted using Cochran’s Q test.

For the *cis*-only analysis, we applied a generalised IVW MR model followed by generalised Egger regression to account for LD structure between correlated SNPs in the *SOST* region and to boost statistical power(*76*). The generalised Egger regression intercept term was used as an indicator of potential directional pleiotropy. For the *cis*, *trans*, and *cis*-only analyses, the above-mentioned Bonferroni corrected P-value threshold of 5.0×10^-3^ was used to control for multiple testing.

#### Bidirectional Mendelian randomization analysis of atherosclerosis-related phenotypes on sclerostin

To investigate the possibility of reverse causality between atherosclerosis-related diseases and risk factors and circulating sclerostin level, we used genetic variants associated with 15 atherosclerosis-related diseases and risk factors as genetic predictors (small vessel disease data has no valid genetic predictors, therefore, we were not able to perform bidirectional MR for this trait; for other genetic predictors, the genetic association data were extracted from relevant GWAS listed in **Supplementary Table 4A**). For this analysis, the circulating sclerostin level from our GWAS meta-analysis was used as the outcome. We applied the same five two-sample MR approaches (IVW, MR-Egger, weighted median, single mode estimator and weighted mode estimator)(*74*)(*17*). In addition, due to correlation between lipids and lipoproteins, we further applied a multivariable MR model(*77*) to estimate the independent effect of each lipid and lipoprotein on sclerostin. For the genetic predictors of these lipids and lipoproteins, see **Supplementary Table 4B** and **4C**. We further estimated the strength of the genetic predictors of the 15 atherosclerosis-related diseases and risk factors using F-statistics. To further validate the directionality of the analysis, we conducted Steiger filtering analysis(*78*) of the four selected sclerostin instruments on the 15 atherosclerosis-related diseases and risk factors.

All MR analyses were conducted using the MendelianRandomization R package(*79*) and TwoSampleMR R package (github.com/MRCIEU/TwoSampleMR v0.5.6)(*80*). Results were plotted as forest plots using code derived from the ggplot2 R package.

## Data Availability

All data produced in the present work are contained in the manuscript

## ACKNOWLEDGEMENTS

We are extremely grateful to all the families who took part in the ALSPAC study, the midwives for their help in recruiting them, and the whole ALSPAC team, which includes interviewers, computer and laboratory technicians, clerical workers, research scientists, volunteers, managers, receptionists and nurses. ALSPAC data collection was supported by the Wellcome Trust (grants WT092830M; WT088806; WT102215/2/13/2), UK Medical Research Council (G1001357), and University of Bristol. The UK Medical Research Council and the Wellcome Trust (ref: 102215/2/13/2) and the University of Bristol provide core support for ALSPAC. GDS works in the Medical Research Council Integrative Epidemiology Unit at the University of Bristol MC_UU_00011/1. The Osteoarthritis Initiative (OAI) is a public-private partnership comprised of five contracts (N01-AR-2–2258; N01-AR-2–2259; N01-AR-2–2260; N01-AR-2–2261; N01-AR-2–2262) funded by the NIH. Sclerostin measurement in the OAI was funded by the Wellcome Trust (ref 20378/Z/16/Z) and analyses were supported by NIH R01AR075356 and NIH P30DK072488. The Trøndelag Health Study (HUNT) is a collaboration between HUNT Research Centre (Faculty of Medicine and Health Sciences, NTNU, Norwegian University of Science and Technology), Trøndelag County Council, Central Norway Regional Health Authority, and the Norwegian Institute of Public Health. The genotyping in HUNT was financed by the National Institutes of Health; University of Michigan; the Research Council of Norway; the Liaison Committee for Education, Research and Innovation in Central Norway; and the Joint Research Committee between St Olavs hospital and the Faculty of Medicine and Health Sciences, NTNU. The genetic investigations of the HUNT Study is a collaboration between researchers from the K.G. Jebsen Center for Genetic Epidemiology, NTNU and the University of Michigan Medical School and the University of Michigan School of Public Health. The K.G. Jebsen Center for Genetic Epidemiology is financed by Stiftelsen Kristian Gerhard Jebsen; Faculty of Medicine and Health Sciences, NTNU, Norway. S.W.v.d.L is funded through EU H2020 TO_AITION (grant number: 848146). We are thankful for the support of the Netherlands CardioVascular Research Initiative of the Netherlands Heart Foundation (CVON 2011/B019 and CVON 2017-20: Generating the best evidence-based pharmaceutical targets for atherosclerosis [GENIUS I&II]), the ERA-CVD program ‘druggable-MI-targets’ (grant number: 01KL1802), and the Leducq Fondation ‘PlaqOmics’. Dr. Rajeev Malhotra was supported by the National Heart, Lung, and Blood Institute (R01HL142809 and R01HL159514), the American Heart Association (18TPA34230025), and the Wild Family Foundation. J.P.K is funded by a National Health and Medical Research Council (Australia) Investigator grant (GNT1177938). Paul S. de Vries and Patricia A. Peyser were supported by National Heart, Lung and Blood Institute (NHLBI) grant number R01HL146860. Infrastructure for the CHARGE Consortium was supported in part by the NHLBI grant R01HL105756.

## REFERENCES

1. E. M. Lewiecki, T. Blicharski, S. Goemaere, K. Lippuner, P. D. Meisner, P. D. Miller, A. Miyauchi, J. Maddox, L. Chen, S. Horlait, A Phase III Randomized Placebo-Controlled Trial to Evaluate Efficacy and Safety of Romosozumab in Men With Osteoporosis. J. Clin. Endocrinol. Metab. 103, 3183–3193 (2018).

2. K. G. Saag, J. Petersen, A. Grauer, Romosozumab versus Alendronate and Fracture Risk in Women with Osteoporosis. N. Engl. J. Med. 378, 195–196 (2018).

3. F. Cosman, D. B. Crittenden, J. D. Adachi, N. Binkley, E. Czerwinski, S. Ferrari, L. C. Hofbauer, E. Lau, E. M. Lewiecki, A. Miyauchi, C. A. F. Zerbini, C. E. Milmont, L. Chen, J. Maddox, P. D. Meisner, C. Libanati, A. Grauer, Romosozumab Treatment in Postmenopausal Women with Osteoporosis. N. Engl. J. Med. 375, 1532–1543 (2016).

4. I. R. Reid, A. M. Horne, B. Mihov, A. Stewart, E. Garratt, S. Bastin, G. D. Gamble, Effects of Zoledronate on Cancer, Cardiac Events, and Mortality in Osteopenic Older Women. J. Bone Miner. Res. 35, 20–27 (2020).

5. S. R. Cummings, L.-Y. Lui, R. Eastell, I. E. Allen, Association Between Drug Treatments for Patients With Osteoporosis and Overall Mortality Rates: A Meta-analysis. JAMA Intern. Med. 179, 1491–1500 (2019).

6. A. De Maré, B. Opdebeeck, E. Neven, P. C. D’Haese, A. Verhulst, Sclerostin Protects Against Vascular Calcification Development in Mice. J. Bone Miner. Res. (2022), doi:10.1002/jbmr.4503.

7. M. Frysz, I. Gergei, H. Scharnagl, G. D. Smith, J. Zheng, D. A. Lawlor, M. Herrmann, W. Maerz, J. H. Tobias, Circulating sclerostin levels are positively related to coronary artery disease severity and related risk factors. J. Bone Miner. Res. (2021), doi:10.1002/jbmr.4467.

8. P. Garnero, E. Sornay-Rendu, F. Munoz, O. Borel, R. D. Chapurlat, Association of serum sclerostin with bone mineral density, bone turnover, steroid and parathyroid hormones, and fracture risk in postmenopausal women: the OFELY study. Osteoporos. Int. 24, 489–494 (2013).

9. G. D. Smith, S. Ebrahim, Data dredging, bias, or confounding. BMJ 325, 1437–1438 (2002).

10. G. Davey Smith, S. Ebrahim, ‘Mendelian randomization’: can genetic epidemiology contribute to understanding environmental determinants of disease? Int. J. Epidemiol. 32, 1–22 (2003).

11. R. C. Richmond, G. Davey Smith, Mendelian Randomization: Concepts and Scope. Cold Spring Harb. Perspect. Med. 12 (2022), doi:10.1101/cshperspect.a040501.

12. J. Bovijn, K. Krebs, C.-Y. Chen, R. Boxall, J. C. Censin, T. Ferreira, S. L. Pulit, C. A. Glastonbury, S. Laber, I. Y. Millwood, K. Lin, L. Li, Z. Chen, L. Milani, G. D. Smith, R. G. Walters, R. Mägi, B. M. Neale, C. M. Lindgren, M. V. Holmes, Evaluating the cardiovascular safety of sclerostin inhibition using evidence from meta-analysis of clinical trials and human genetics. Sci. Transl. Med. 12 (2020), doi:10.1126/scitranslmed.aay6570.

13. Committee for Medicinal Products for Human Use (CHMP), Assessment report for romosozumab. European Medicines Agent (2019) (available at https://www.ema.europa.eu/en/documents/assessment-report/evenity-epar-public-assessment-report_en.pdf).

14. G. Holdsworth, J. R. Staley, P. Hall, I. van Koeverden, C. Vangjeli, R. Okoye, R. W. Boyce, J. R. Turk, M. Armstrong, A. Wolfreys, G. Pasterkamp, Sclerostin Downregulation Globally by Naturally Occurring Genetic Variants, or Locally in Atherosclerotic Plaques, Does Not Associate With Cardiovascular Events in Humans. J. Bone Miner. Res. (2021), doi:10.1002/jbmr.4287.

15. J. Zheng, W. Maerz, I. Gergei, M. Kleber, C. Drechsler, C. Wanner, V. Brandenburg, S. Reppe, K. Gautvik, C. Medina-Gomez, E. Shevroja, A. Gilly, Y.-C. Park, G. Dedoussis, E. Zeggini, M. Lorentzon, P. Henning, U. Lerne, K. Nilsson, S. Moverare-Skrtic, D. Baird, L. Falk, A. Groom, T. Capellini, E. Grundberg, M. Nethander, C. Ohlsson, G. D. Smith, J. Tobias, Genome-wide mapping identifies beta-1,4-N-acetyl-galactosaminyl-transferase as a novel determinant of sclerostin levels and bone mineral density. J. Bone Miner. Res. (2018).

16. G. Davey Smith, G. Hemani, Mendelian randomization: genetic anchors for causal inference in epidemiological studies. Hum. Mol. Genet. 23, R89–98 (2014).

17. E. Sanderson, M. M. Glymour, M. V. Holmes, H. Kang, J. Morrison, M. R. Munafò, T. Palmer, C. Mary Schooling, C. Wallace, Q. Zhao, G. D. Smith, Mendelian randomization. Nature Reviews Methods Primers 2, 1–1 (2022).

18. M. V. Holmes, T. G. Richardson, B. A. Ference, N. M. Davies, G. Davey Smith, Integrating genomics with biomarkers and therapeutic targets to invigorate cardiovascular drug development. Nat. Rev. Cardiol. 18, 435–453 (2021).

19. M. Pietzner, E. Wheeler, J. Carrasco-Zanini, A. Cortes, M. Koprulu, M. A. Wörheide, E. Oerton, J. Cook, I. D. Stewart, N. D. Kerrison, J. Luan, J. Raffler, M. Arnold, W. Arlt, S. O’Rahilly, G. Kastenmüller, E. R. Gamazon, A. D. Hingorani, R. A. Scott, N. J. Wareham, C. Langenberg, Mapping the proteo-genomic convergence of human diseases. Science, eabj1541 (2021).

20. E. Di Angelantonio, S. G. Thompson, S. Kaptoge, C. Moore, M. Walker, J. Armitage, W. H. Ouwehand, D. J. Roberts, J. Danesh, INTERVAL Trial Group, Efficiency and safety of varying the frequency of whole blood donation (INTERVAL): a randomised trial of 45 000 donors. Lancet 390, 2360–2371 (2017).

21. S. Krokstad, A. Langhammer, K. Hveem, T. L. Holmen, K. Midthjell, T. R. Stene, G. Bratberg, J. Heggland, J. Holmen, Cohort Profile: the HUNT Study, Norway. Int. J. Epidemiol. 42, 968–977 (2013).

22. B. O. Åsvold, A. Langhammer, T. A. Rehn, G. Kjelvik, T. V. Grøntvedt, E. P. Sørgjerd, J. S. Fenstad, O. Holmen, M. C. Stuifbergen, S. A. A. Vikjord, B. M. Brumpton, H. K. Skjellegrind, P. Thingstad, E. R. Sund, G. Selbæk, P. J. Mork, V. Rangul, K. Hveem, M. Næss, S. Krokstad, Cohort profile update: The HUNT study, Norway. bioRxiv (2021), doi:10.1101/2021.10.12.21264858.

23. B. M. Brumpton, S. Graham, I. Surakka, A. H. Skogholt, M. Løset, L. G. Fritsche, B. Wolford, W. Zhou, J. B. Nielsen, O. L. Holmen, M. E. Gabrielsen, L. Thomas, L. Bhatta, H. Rasheed, H. Zhang, H. M. Kang, W. Hornsby, M. R. Moksnes, E. Coward, M. Melbye, G. F. Giskeødegård, J. Fenstad, S. Krokstad, M. Næss, A. Langhammer, M. Boehnke, G. R. Abecasis, B. O. Åsvold, K. Hveem, C. J. Willer, The HUNT Study: a population-based cohort for genetic research. bioRxiv (2021), doi:10.1101/2021.12.23.21268305.

24. G. Lester, The Osteoarthritis Initiative: A NIH Public-Private Partnership. HSS J. 8, 62–63 (2012).

25. L. K. Conlin, B. D. Thiel, C. G. Bonnemann, L. Medne, L. M. Ernst, E. H. Zackai, M. A. Deardorff, I. D. Krantz, H. Hakonarson, N. B. Spinner, Mechanisms of mosaicism, chimerism and uniparental disomy identified by single nucleotide polymorphism array analysis. Hum. Mol. Genet. 19, 1263–1275 (2010).

26. D. A. Peiffer, J. M. Le, F. J. Steemers, W. Chang, T. Jenniges, F. Garcia, K. Haden, J. Li, C. A. Shaw, J. Belmont, S. W. Cheung, R. M. Shen, D. L. Barker, K. L. Gunderson, High-resolution genomic profiling of chromosomal aberrations using Infinium whole-genome genotyping. Genome Res. 16, 1136–1148 (2006).

27. C. C. Laurie, K. F. Doheny, D. B. Mirel, E. W. Pugh, L. J. Bierut, T. Bhangale, F. Boehm, N. E. Caporaso, M. C. Cornelis, H. J. Edenberg, S. B. Gabriel, E. L. Harris, F. B. Hu, K. B. Jacobs, P. Kraft, M. T. Landi, T. Lumley, T. A. Manolio, C. McHugh, I. Painter, J. Paschall, J. P. Rice, K. M. Rice, X. Zheng, B. S. Weir, GENEVA Investigators, Quality control and quality assurance in genotypic data for genome-wide association studies. Genet. Epidemiol. 34, 591–602 (2010).

28. B. R. Winkelmann, W. März, B. O. Boehm, R. Zotz, J. Hager, P. Hellstern, J. Senges, LURIC Study Group (LUdwigshafen RIsk and Cardiovascular Health), Rationale and design of the LURIC study--a resource for functional genomics, pharmacogenomics and long-term prognosis of cardiovascular disease. Pharmacogenomics 2, S1–73 (2001).

29. T. F. M. Andlauer, D. Buck, G. Antony, A. Bayas, L. Bechmann, A. Berthele, A. Chan, C. Gasperi, R. Gold, C. Graetz, J. Haas, M. Hecker, C. Infante-Duarte, M. Knop, T. Kümpfel, V. Limmroth, R. A. Linker, V. Loleit, F. Luessi, S. G. Meuth, M. Mühlau, S. Nischwitz, F. Paul, M. Pütz, T. Ruck, A. Salmen, M. Stangel, J.-P. Stellmann, K. H. Stürner, B. Tackenberg, F. Then Bergh, H. Tumani, C. Warnke, F. Weber, H. Wiendl, B. Wildemann, U. K. Zettl, U. Ziemann, F. Zipp, J. Arloth, P. Weber, M. Radivojkov-Blagojevic, M. O. Scheinhardt, T. Dankowski, T. Bettecken, P. Lichtner, D. Czamara, T. Carrillo-Roa, E. B. Binder, K. Berger, L. Bertram, A. Franke, C. Gieger, S. Herms, G. Homuth, M. Ising, K.-H. Jöckel, T. Kacprowski, S. Kloiber, M. Laudes, W. Lieb, C. M. Lill, S. Lucae, T. Meitinger, S. Moebus, M. Müller-Nurasyid, M. M. Nöthen, A. Petersmann, R. Rawal, U. Schminke, K. Strauch, H. Völzke, M. Waldenberger, J. Wellmann, E. Porcu, A. Mulas, M. Pitzalis, C. Sidore, I. Zara, F. Cucca, M. Zoledziewska, A. Ziegler, B. Hemmer, B. Müller-Myhsok, Novel multiple sclerosis susceptibility loci implicated in epigenetic regulation. Sci Adv 2, e1501678 (2016).

30. F. Mihalache, A. Höblinger, F. Grünhage, M. Krawczyk, B. C. Gärtner, M. Acalovschi, T. Sauerbruch, F. Lammert, V. Zimmer, Heterozygosity for the alpha1-antitrypsin Z allele may confer genetic risk of cholangiocarcinoma. Aliment. Pharmacol. Ther. 33, 389–394 (2011).

31. J. P. Kemp, C. Medina-Gomez, K. Estrada, B. St Pourcain, D. H. M. Heppe, N. M. Warrington, L. Oei, S. M. Ring, C. J. Kruithof, N. J. Timpson, L. E. Wolber, S. Reppe, K. Gautvik, E. Grundberg, B. Ge, B. van der Eerden, J. van de Peppel, M. A. Hibbs, C. L. Ackert-Bicknell, K. Choi, D. L. Koller, M. J. Econs, F. M. K. Williams, T. Foroud, M. C. Zillikens, C. Ohlsson, A. Hofman, A. G. Uitterlinden, G. Davey Smith, V. W. V. Jaddoe, J. H. Tobias, F. Rivadeneira, D. M. Evans, Phenotypic dissection of bone mineral density reveals skeletal site specificity and facilitates the identification of novel loci in the genetic regulation of bone mass attainment. PLoS Genet. 10, e1004423 (2014).

32. F. Rivadeneira, U. Styrkársdottir, K. Estrada, B. V. Halldórsson, Y.-H. Hsu, J. B. Richards, M. C. Zillikens, F. K. Kavvoura, N. Amin, Y. S. Aulchenko, L. A. Cupples, P. Deloukas, S. Demissie, E. Grundberg, A. Hofman, A. Kong, D. Karasik, J. B. van Meurs, B. Oostra, T. Pastinen, H. A. P. Pols, G. Sigurdsson, N. Soranzo, G. Thorleifsson, U. Thorsteinsdottir, F. M. K. Williams, S. G. Wilson, Y. Zhou, S. H. Ralston, C. M. van Duijn, T. Spector, D. P. Kiel, K. Stefansson, J. P. A. Ioannidis, A. G. Uitterlinden, Genetic Factors for Osteoporosis (GEFOS) Consortium, Twenty bone-mineral-density loci identified by large-scale meta-analysis of genome-wide association studies. Nat. Genet. 41, 1199–1206 (2009).

33. R. Jemtland, M. Holden, S. Reppe, O. K. Olstad, F. P. Reinholt, V. T. Gautvik, H. Refvem, A. Frigessi, B. Houston, K. M. Gautvik, Molecular disease map of bone characterizing the postmenopausal osteoporosis phenotype. J. Bone Miner. Res. 26, 1793–1801 (2011).

34. J. Ernst, M. Kellis, ChromHMM: automating chromatin-state discovery and characterization. Nat. Methods 9, 215–216 (2012).

35. A. P. Boyle, E. L. Hong, M. Hariharan, Y. Cheng, M. A. Schaub, M. Kasowski, K. J. Karczewski, J. Park, B. C. Hitz, S. Weng, J. M. Cherry, M. Snyder, Annotation of functional variation in personal genomes using RegulomeDB. Genome Res. 22, 1790–1797 (2012).

36. GTEx Consortium, The GTEx Consortium atlas of genetic regulatory effects across human tissues. Science 369, 1318–1330 (2020).

37. J. Støy, U. Kampmann, A. Mengel, N. E. Magnusson, N. Jessen, N. Grarup, J. Rungby, H. Stødkilde-Jørgensen, I. Brandslund, C. Christensen, T. Hansen, O. Pedersen, N. Møller, Reduced CD300LG mRNA tissue expression, increased intramyocellular lipid content and impaired glucose metabolism in healthy male carriers of Arg82Cys in CD300LG: a novel genometabolic cross-link between CD300LG and common metabolic phenotypes. BMJ Open Diabetes Research and Care 3, e000095 (2015).

38. N. J. Timpson, B. G. Nordestgaard, R. M. Harbord, J. Zacho, T. M. Frayling, A. Tybjærg-Hansen, G. D. Smith, C-reactive protein levels and body mass index: elucidating direction of causation through reciprocal Mendelian randomization. Int. J. Obes. 35, 300–308 (2011).

39. A. R. van Rosendael, I. J. van den Hoogen, U. Gianni, X. Ma, S. W. Tantawy, A. M. Bax, Y. Lu, D. Andreini, M. H. Al-Mallah, M. J. Budoff, F. Cademartiri, K. Chinnaiyan, J. H. Choi, E. Conte, H. Marques, P. de Araújo Gonçalves, I. Gottlieb, M. Hadamitzky, J. A. Leipsic, E. Maffei, G. Pontone, S. Shin, Y.-J. Kim, B. K. Lee, E. J. Chun, J. M. Sung, S.-E. Lee, R. Virmani, H. Samady, Y. Sato, P. H. Stone, D. S. Berman, J. Narula, R. Blankstein, J. K. Min, F. Y. Lin, L. J. Shaw, J. J. Bax, H.-J. Chang, Association of Statin Treatment With Progression of Coronary Atherosclerotic Plaque Composition. JAMA Cardiol (2021), doi:10.1001/jamacardio.2021.3055.

40. G. S. Gulsin, A. J. Moss, Coronary artery calcium paradox and physical activity. Heart 107, 1686–1687 (2021).

41. K.-C. Sung, Y. S. Hong, J.-Y. Lee, S.-J. Lee, Y. Chang, S. Ryu, D. Zhao, J. Cho, E. Guallar, J. A. C. Lima, Physical activity and the progression of coronary artery calcification. Heart (2021), doi:10.1136/heartjnl-2021-319346.

42. G. L. Galea, L. E. Lanyon, J. S. Price, Sclerostin’s role in bone’s adaptive response to mechanical loading. Bone 96, 38–44 (2017).

43. R. Koos, V. Brandenburg, A. H. Mahnken, R. Schneider, G. Dohmen, R. Autschbach, N. Marx, R. Kramann, Sclerostin as a potential novel biomarker for aortic valve calcification: an in-vivo and ex-vivo study. J. Heart Valve Dis. 22, 317–325 (2013).

44. D. Zhu, N. C. W. Mackenzie, J. L. Millán, C. Farquharson, V. E. MacRae, The appearance and modulation of osteocyte marker expression during calcification of vascular smooth muscle cells. PLoS One 6, e19595 (2011).

45. X. Li, Y. Zhang, H. Kang, W. Liu, P. Liu, J. Zhang, S. E. Harris, D. Wu, Sclerostin binds to LRP5/6 and antagonizes canonical Wnt signaling. J. Biol. Chem. 280, 19883–19887 (2005).

46. A. Catalano, F. Bellone, N. Morabito, F. Corica, Sclerostin and Vascular Pathophysiology. Int. J. Mol. Sci. 21 (2020), doi:10.3390/ijms21134779.

47. S. Y. Lim, M. B. Bolster, Profile of romosozumab and its potential in the management of osteoporosis. Drug Des. Devel. Ther. 11, 1221–1231 (2017).

48. W. Gan, R. J. Clarke, A. Mahajan, B. Kulohoma, H. Kitajima, N. R. Robertson, N. W. Rayner, R. G. Walters, M. V. Holmes, Z. Chen, M. I. McCarthy, Bone mineral density and risk of type 2 diabetes and coronary heart disease: A Mendelian randomization study. Wellcome Open Res 2, 68 (2017).

49. Committee for Medicinal Products for Human Use (CHMP), Recommendation to restrict the use of Protelos/Osseor (strontium ranelate). European Medicines Agent (2013) (available at https://www.ema.europa.eu/en/documents/press-release/recommendation-restrict-use-protelos/osseor-strontium-ranelate_en.pdf).

50. O. M. E. Albagha, S. E. Wani, M. R. Visconti, N. Alonso, K. Goodman, M. L. Brandi, T. Cundy, P. Y. J. Chung, R. Dargie, J.-P. Devogelaer, A. Falchetti, W. D. Fraser, L. Gennari, F. Gianfrancesco, M. J. Hooper, W. Van Hul, G. Isaia, G. C. Nicholson, R. Nuti, S. Papapoulos, J. del P. Montes, T. Ratajczak, S. L. Rea, D. Rendina, R. Gonzalez-Sarmiento, M. Di Stefano, L. C. Ward, J. P. Walsh, S. H. Ralston, Genetic Determinants of Paget’s Disease (GDPD) Consortium, Genome-wide association identifies three new susceptibility loci for Paget’s disease of bone. Nat. Genet. 43, 685–689 (2011).

51. I. Tachmazidou, D. Süveges, J. L. Min, G. R. S. Ritchie, J. Steinberg, K. Walter, V. Iotchkova, J. Schwartzentruber, J. Huang, Y. Memari, S. McCarthy, A. A. Crawford, C. Bombieri, M. Cocca, A.-E. Farmaki, T. R. Gaunt, P. Jousilahti, M. N. Kooijman, B. Lehne, G. Malerba, S. Männistö, A. Matchan, C. Medina-Gomez, S. J. Metrustry, A. Nag, I. Ntalla, L. Paternoster, N. W. Rayner, C. Sala, W. R. Scott, H. A. Shihab, L. Southam, B. St Pourcain, M. Traglia, K. Trajanoska, G. Zaza, W. Zhang, M. S. Artigas, N. Bansal, M. Benn, Z. Chen, P. Danecek, W.-Y. Lin, A. Locke, J. Luan, A. K. Manning, A. Mulas, C. Sidore, A. Tybjaerg-Hansen, A. Varbo, M. Zoledziewska, C. Finan, K. Hatzikotoulas, A. E. Hendricks, J. P. Kemp, A. Moayyeri, K. Panoutsopoulou, M. Szpak, S. G. Wilson, M. Boehnke, F. Cucca, E. Di Angelantonio, C. Langenberg, C. Lindgren, M. I. McCarthy, A. P. Morris, B. G. Nordestgaard, R. A. Scott, M. D. Tobin, N. J. Wareham, SpiroMeta Consortium, GoT2D Consortium, P. Burton, J. C. Chambers, G. D. Smith, G. Dedoussis, J. F. Felix, O. H. Franco, G. Gambaro, P. Gasparini, C. J. Hammond, A. Hofman, V. W. V. Jaddoe, M. Kleber, J. S. Kooner, M. Perola, C. Relton, S. M. Ring, F. Rivadeneira, V. Salomaa, T. D. Spector, O. Stegle, D. Toniolo, A. G. Uitterlinden, arcOGEN Consortium, Understanding Society Scientific Group, UK10K Consortium, I. Barroso, C. M. T. Greenwood, J. R. B. Perry, B. R. Walker, A. S. Butterworth, Y. Xue, R. Durbin, K. S. Small, N. Soranzo, N. J. Timpson, E. Zeggini, Whole-Genome Sequencing Coupled to Imputation Discovers Genetic Signals for Anthropometric Traits. Am. J. Hum. Genet. 100, 865–884 (2017).

52. J. Mbatchou, L. Barnard, J. Backman, A. Marcketta, J. A. Kosmicki, A. Ziyatdinov, C. Benner, C. O’Dushlaine, M. Barber, B. Boutkov, L. Habegger, M. Ferreira, A. Baras, J. Reid, G. Abecasis, E. Maxwell, J. Marchini, Computationally efficient whole-genome regression for quantitative and binary traits. Nat. Genet. 53, 1097–1103 (2021).

53. C. C. Chang, C. C. Chow, L. C. Tellier, S. Vattikuti, S. M. Purcell, J. J. Lee, Second-generation PLINK: rising to the challenge of larger and richer datasets. Gigascience 4, 7 (2015).

54. T. W. Winkler, F. R. Day, D. C. Croteau-Chonka, A. R. Wood, A. E. Locke, R. Mägi, T. Ferreira, T. Fall, M. Graff, A. E. Justice, J. Luan, S. Gustafsson, J. C. Randall, S. Vedantam, T. Workalemahu, T. O. Kilpeläinen, A. Scherag, T. Esko, Z. Kutalik, I. M. Heid, R. J. F. Loos, Genetic Investigation of Anthropometric Traits (GIANT) Consortium, Quality control and conduct of genome-wide association meta-analyses. Nat. Protoc. 9, 1192–1212 (2014).

55. C. J. Willer, Y. Li, G. R. Abecasis, METAL: fast and efficient meta-analysis of genomewide association scans. Bioinformatics 26, 2190–2191 (2010).

56. Y. Li, C. J. Willer, J. Ding, P. Scheet, G. R. Abecasis, MaCH: using sequence and genotype data to estimate haplotypes and unobserved genotypes. Genet. Epidemiol. 34, 816–834 (2010).

57. B. Howie, C. Fuchsberger, M. Stephens, J. Marchini, G. R. Abecasis, Fast and accurate genotype imputation in genome-wide association studies through pre-phasing. Nat. Genet. 44, 955–959 (2012).

58. R. Mägi, A. P. Morris, GWAMA: software for genome-wide association meta-analysis. BMC Bioinformatics 11, 288 (2010).

59. J. Yang, T. Ferreira, A. P. Morris, S. E. Medland, Genetic Investigation of ANthropometric Traits (GIANT) Consortium, DIAbetes Genetics Replication And Meta-analysis (DIAGRAM) Consortium, P. A. F. Madden, A. C. Heath, N. G. Martin, G. W. Montgomery, M. N. Weedon, R. J. Loos, T. M. Frayling, M. I. McCarthy, J. N. Hirschhorn, M. E. Goddard, P. M. Visscher, Conditional and joint multiple-SNP analysis of GWAS summary statistics identifies additional variants influencing complex traits. Nat. Genet. 44, 369–75, S1-3 (2012).

60. C. Giambartolomei, D. Vukcevic, E. E. Schadt, L. Franke, A. D. Hingorani, C. Wallace, V. Plagnol, Bayesian test for colocalisation between pairs of genetic association studies using summary statistics. PLoS Genet. 10, e1004383 (2014).

61. K. Watanabe, E. Taskesen, A. van Bochoven, D. Posthuma, Functional mapping and annotation of genetic associations with FUMA. Nat. Commun. 8, 1–11 (2017).

62. S. Koplev, M. Seldin, K. Sukhavasi, R. Ermel, S. Pang, L. Zeng, S. Bankier, A. Di Narzo, H. Cheng, V. Meda, A. Ma, H. Talukdar, A. Cohain, L. Amadori, C. Argmann, S. M. Houten, O. Franzén, G. Mocci, O. A. Meelu, K. Ishikawa, C. Whatling, A. Jain, R. K. Jain, L.-M. Gan, C. Giannarelli, P. Roussos, K. Hao, H. Schunkert, T. Michoel, A. Ruusalepp, E. E. Schadt, J. C. Kovacic, A. J. Lusis, J. L. M. Björkegren, A mechanistic framework for cardiometabolic and coronary artery diseases. Nature Cardiovascular Research 1, 85–100 (2022).

63. B. K. Bulik-Sullivan, P.-R. Loh, H. K. Finucane, S. Ripke, J. Yang, Schizophrenia Working Group of the Psychiatric Genomics Consortium, N. Patterson, M. J. Daly, A. L. Price, B. M. Neale, LD Score regression distinguishes confounding from polygenicity in genome-wide association studies. Nat. Genet. 47, 291–295 (2015).

64. J. Zheng, A. M. Erzurumluoglu, B. L. Elsworth, J. P. Kemp, L. Howe, P. C. Haycock, G. Hemani, K. Tansey, C. Laurin, Early Genetics and Lifecourse Epidemiology (EAGLE) Eczema Consortium, B. S. Pourcain, N. M. Warrington, H. K. Finucane, A. L. Price, B. K. Bulik-Sullivan, V. Anttila, L. Paternoster, T. R. Gaunt, D. M. Evans, B. M. Neale, LD Hub: a centralized database and web interface to perform LD score regression that maximizes the potential of summary level GWAS data for SNP heritability and genetic correlation analysis. Bioinformatics 33, 272–279 (2017).

65. D. A. Lawlor, R. M. Harbord, J. A. C. Sterne, N. Timpson, G. Davey Smith, Mendelian randomization: using genes as instruments for making causal inferences in epidemiology. Stat. Med. 27, 1133–1163 (2008).

66. J. Zheng, Richardson T.R., Millard L., Hemani G., Raistrick C., Vilhjalmsson B., Haycock P.C., Gaunt T.R., PhenoSpD: an integrated toolkit for phenotypic correlation estimation and multiple testing correction using GWAS summary statistics. GigaScience (2018) doi:10.1101/148627.

67. A. Mahajan, D. Taliun, M. Thurner, N. R. Robertson, J. M. Torres, N. W. Rayner, A. J. Payne, V. Steinthorsdottir, R. A. Scott, N. Grarup, J. P. Cook, E. M. Schmidt, M. Wuttke, C. Sarnowski, R. Mägi, J. Nano, C. Gieger, S. Trompet, C. Lecoeur, M. H. Preuss, B. P. Prins, X. Guo, L. F. Bielak, J. E. Below, D. W. Bowden, J. C. Chambers, Y. J. Kim, M. C. Y. Ng, L. E. Petty, X. Sim, W. Zhang, A. J. Bennett, J. Bork-Jensen, C. M. Brummett, M. Canouil, K.-U. Ec Kardt, K. Fischer, S. L. R. Kardia, F. Kronenberg, K. Läll, C.-T. Liu, A. E. Locke, J. Luan, I. Ntalla, V. Nylander, S. Schönherr, C. Schurmann, L. Yengo, E. P. Bottinger, I. Brandslund, C. Christensen, G. Dedoussis, J. C. Florez, I. Ford, O. H. Franco, T. M. Frayling, V. Giedraitis, S. Hackinger, A. T. Hattersley, C. Herder, M. A. Ikram, M. Ingelsson, M. E. Jørgensen, T. Jørgensen, J. Kriebel, J. Kuusisto, S. Ligthart, C. M. Lindgren, A. Linneberg, V. Lyssenko, V. Mamakou, T. Meitinger, K. L. Mohlke, A. D. Morris, G. Nadkarni, J. S. Pankow, A. Peters, N. Sattar, A. Stančáková, K. Strauch, K. D. Taylor, B. Thorand, G. Thorleifsson, U. Thorsteinsdottir, J. Tuomilehto, D. R. Witte, J. Dupuis, P. A. Peyser, E. Zeggini, R. J. F. Loos, P. Froguel, E. Ingelsson, L. Lind, L. Groop, M. Laakso, F. S. Collins, J. W. Jukema, C. N. A. Palmer, H. Grallert, A. Metspalu, A. Dehghan, A. Köttgen, G. R. Abecasis, J. B. Meigs, J. I. Rotter, J. Marchini, O. Pedersen, T. Hansen, C. Langenberg, N. J. Wareham, K. Stefansson, A. L. Gloyn, A. P. Morris, M. Boehnke, M. I. McCarthy, Fine-mapping type 2 diabetes loci to single-variant resolution using high-density imputation and islet-specific epigenome maps. Nat. Genet. 50, 1505–1513 (2018).

68. R. Malik, M. Traylor, S. L. Pulit, S. Bevan, J. C. Hopewell, E. G. Holliday, W. Zhao, P. Abrantes, P. Amouyel, J. R. Attia, T. W. K. Battey, K. Berger, G. B. Boncoraglio, G. Chauhan, Y.-C. Cheng, W.-M. Chen, R. Clarke, I. Cotlarciuc, S. Debette, G. J. Falcone, J. M. Ferro, D. M. Gamble, A. Ilinca, S. J. Kittner, C. E. Kourkoulis, R. Lemmens, C. R. Levi, P. Lichtner, A. Lindgren, J. Liu, J. F. Meschia, B. D. Mitchell, S. A. Oliveira, J. Pera, A. P. Reiner, P. M. Rothwell, P. Sharma, A. Slowik, C. L. M. Sudlow, T. Tatlisumak, V. Thijs, A. M. Vicente, D. Woo, S. Seshadri, D. Saleheen, J. Rosand, H. S. Markus, B. B. Worrall, M. Dichgans, ISGC Analysis Group, METASTROKE collaboration, Wellcome Trust Case Control Consortium 2 (WTCCC2), NINDS Stroke Genetics Network (SiGN), Low-frequency and common genetic variation in ischemic stroke: The METASTROKE collaboration. Neurology 86, 1217–1226 (2016).

69. P. van der Harst, N. Verweij, Identification of 64 Novel Genetic Loci Provides an Expanded View on the Genetic Architecture of Coronary Artery Disease. Circ. Res. 122, 433–443 (2018).

70. J. A. Hartiala, Y. Han, Q. Jia, J. R. Hilser, P. Huang, J. Gukasyan, W. S. Schwartzman, Z. Cai, S. Biswas, D.-A. Trégouët, N. L. Smith, INVENT Consortium, CHARGE Consortium Hemostasis Working Group, GENIUS-CHD Consortium, M. Seldin, C. Pan, M. Mehrabian, A. J. Lusis, P. Bazeley, Y. V. Sun, C. Liu, A. A. Quyyumi, M. Scholz, J. Thiery, G. E. Delgado, M. E. Kleber, W. März, L. J. Howe, F. W. Asselbergs, M. van Vugt, G. J. Vlachojannis, R. S. Patel, L.-P. Lyytikäinen, M. Kähönen, T. Lehtimäki, T. V. M. Nieminen, P. Kuukasjärvi, J. O. Laurikka, X. Chang, C.-K. Heng, R. Jiang, W. E. Kraus, E. R. Hauser, J. F. Ferguson, M. P. Reilly, K. Ito, S. Koyama, Y. Kamatani, I. Komuro, Biobank Japan, L. K. Stolze, C. E. Romanoski, M. D. Khan, A. W. Turner, C. L. Miller, R. Aherrahrou, M. Civelek, L. Ma, J. L. M. Björkegren, S. R. Kumar, W. H. W. Tang, S. L. Hazen, H. Allayee, Genome-wide analysis identifies novel susceptibility loci for myocardial infarction. Eur. Heart J. 42, 919–933 (2021).

71. M. Kavousi, M. M. Bos, H. J. Barnes, C. L. Lino Cardenas, D. Wong, C. J. O’Donnell, L. F. Bielak, P. A. Peyser, R. Malhotra, S. W. van der Laan, C. L. Miller, CHARGE Subclinical and Clinical Atherosclerosis Working Group, Multi-ancestry genome-wide analysis identifies effector genes and druggable pathways for coronary artery calcification (2022), doi:10.1101/2022.05.02.22273844.

72. J. P. Kemp, J. A. Morris, C. Medina-Gomez, V. Forgetta, N. M. Warrington, S. E. Youlten, J. Zheng, C. L. Gregson, E. Grundberg, K. Trajanoska, J. G. Logan, A. S. Pollard, P. C. Sparkes, E. J. Ghirardello, R. Allen, V. D. Leitch, N. C. Butterfield, D. Komla-Ebri, A.-T. Adoum, K. F. Curry, J. K. White, F. Kussy, K. M. Greenlaw, C. Xu, N. C. Harvey, C. Cooper, D. J. Adams, C. M. T. Greenwood, M. T. Maurano, S. Kaptoge, F. Rivadeneira, J. H. Tobias, P. I. Croucher, C. L. Ackert-Bicknell, J. H. D. Bassett, G. R. Williams, J. B. Richards, D. M. Evans, Identification of 153 new loci associated with heel bone mineral density and functional involvement of GPC6 in osteoporosis. Nat. Genet. 49, 1468–1475 (2017).

73. R. Malhotra, A. C. Mauer, C. L. Lino Cardenas, X. Guo, J. Yao, X. Zhang, F. Wunderer, A. V. Smith, Q. Wong, S. Pechlivanis, S.-J. Hwang, J. Wang, L. Lu, C. J. Nicholson, G. Shelton, M. D. Buswell, H. J. Barnes, H. H. Sigurslid, C. Slocum, C. O. Rourke, D. K. Rhee, A. Bagchi, S. U. Nigwekar, E. S. Buys, C. Y. Campbell, T. Harris, M. Budoff, M. H. Criqui, J. I. Rotter, A. D. Johnson, C. Song, N. Franceschini, S. Debette, U. Hoffmann, H. Kälsch, M. M. Nöthen, S. Sigurdsson, B. I. Freedman, D. W. Bowden, K.-H. Jöckel, S. Moebus, R. Erbel, M. F. Feitosa, V. Gudnason, G. Thanassoulis, W. M. Zapol, M. E. Lindsay, D. B. Bloch, W. S. Post, C. J. O’Donnell, HDAC9 is implicated in atherosclerotic aortic calcification and affects vascular smooth muscle cell phenotype. Nat. Genet. 51, 1580–1587 (2019).

74. S. Burgess, A. Butterworth, S. G. Thompson, Mendelian randomization analysis with multiple genetic variants using summarized data. Genet. Epidemiol. 37, 658–665 (2013).

75. J. Bowden, F. Del Greco M, C. Minelli, G. Davey Smith, N. A. Sheehan, J. R. Thompson, Assessing the suitability of summary data for two-sample Mendelian randomization analyses using MR-Egger regression: the role of the I 2 statistic. Int. J. Epidemiol. 45, 1961–1974 (2016).

76. A. Gkatzionis, S. Burgess, P. J. Newcombe, Statistical Methods for cis-Mendelian Randomization. arXiv (2021) (available at http://arxiv.org/abs/2101.04081).

77. S. Burgess, S. G. Thompson, Multivariable Mendelian randomization: the use of pleiotropic genetic variants to estimate causal effects. Am. J. Epidemiol. 181, 251–260 (2015).

78. G. Hemani, K. Tilling, G. Davey Smith, Orienting the causal relationship between imprecisely measured traits using GWAS summary data. PLoS Genet. 13, e1007081 (2017).

79. O. O. Yavorska, S. Burgess, MendelianRandomization: an R package for performing Mendelian randomization analyses using summarized data. Int. J. Epidemiol. (2017), doi:10.1093/ije/dyx034.

80. G. Hemani, J. Zheng, B. Elsworth, K. H. Wade, V. Haberland, D. Baird, C. Laurin, S. Burgess, J. Bowden, R. Langdon, V. Y. Tan, J. Yarmolinsky, H. A. Shihab, N. J. Timpson, D. M. Evans, C. Relton, R. M. Martin, G. Davey Smith, T. R. Gaunt, P. C. Haycock, The MR-Base platform supports systematic causal inference across the human phenome. Elife 7 (2018), doi:10.7554/eLife.34408.

