## Supplementary Figures and notes for "Lowering of circulating sclerostin may increase risk of atherosclerosis and its risk factors: evidence from a genome-wide association meta-analysis followed by Mendelian randomization"

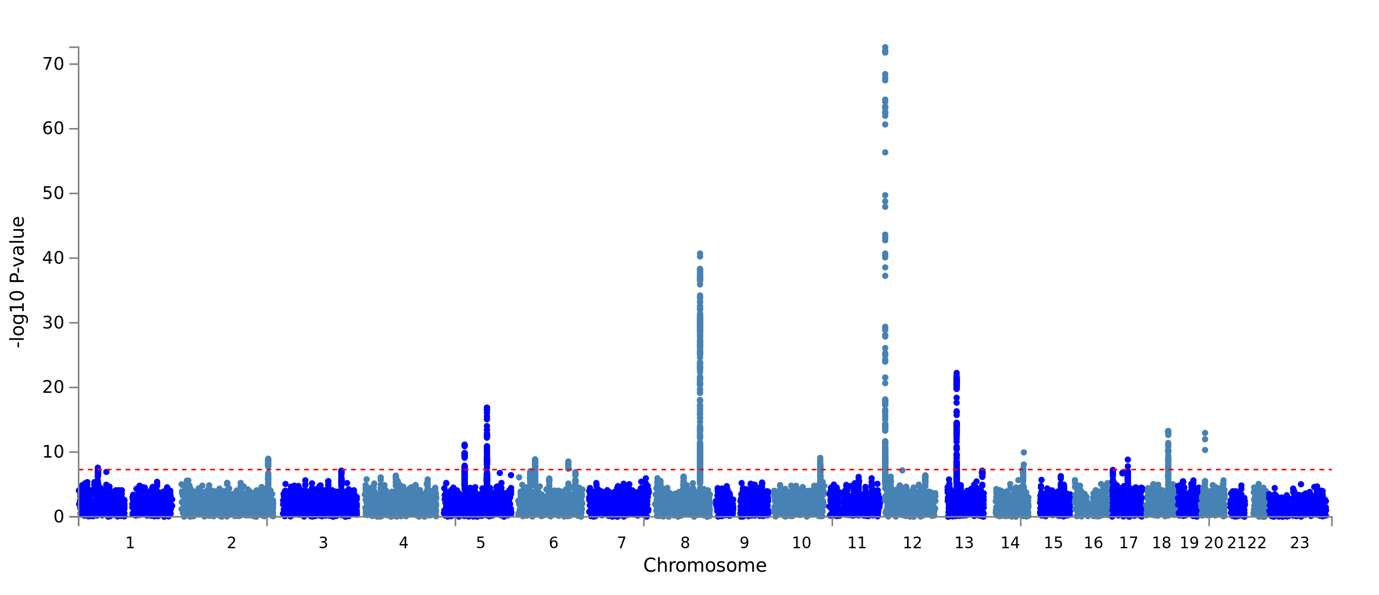

**Supplementary Figure 1**. Manhattan plot of association results from the fixed-effect meta-analysis of sclerostin.

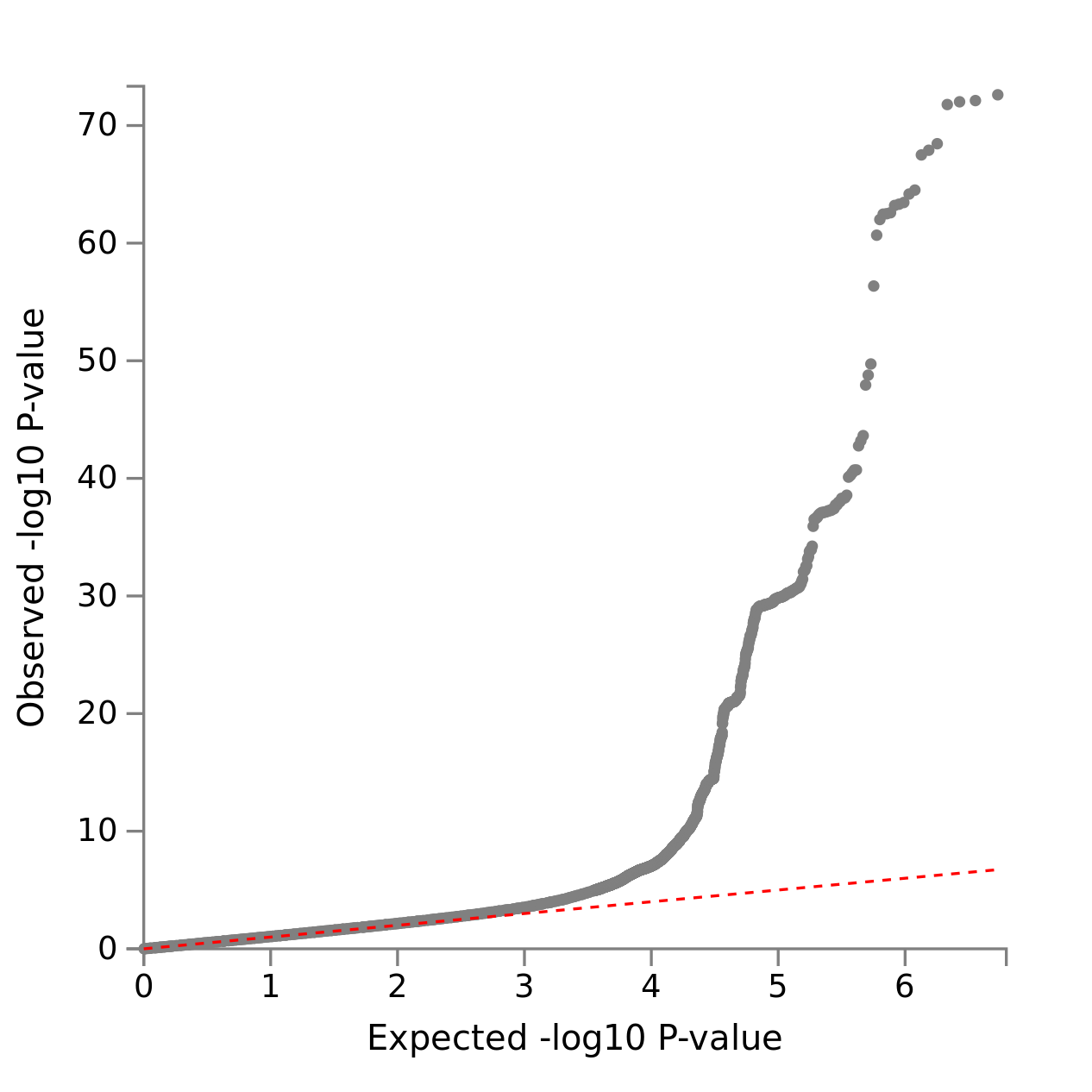

**Supplementary Figure 2**. QQ plot of association results from the fixed-effect meta-analysis of sclerostin.

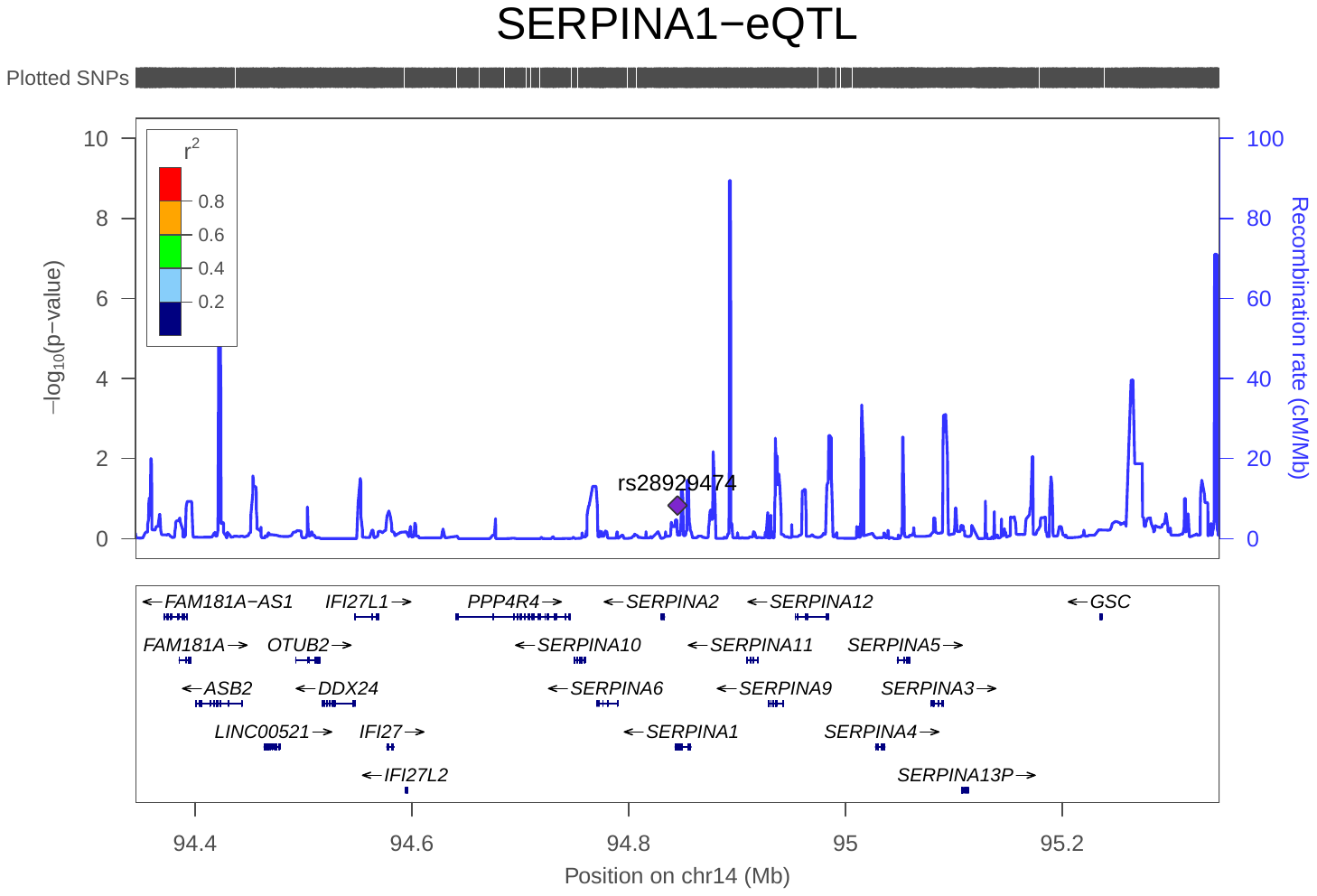

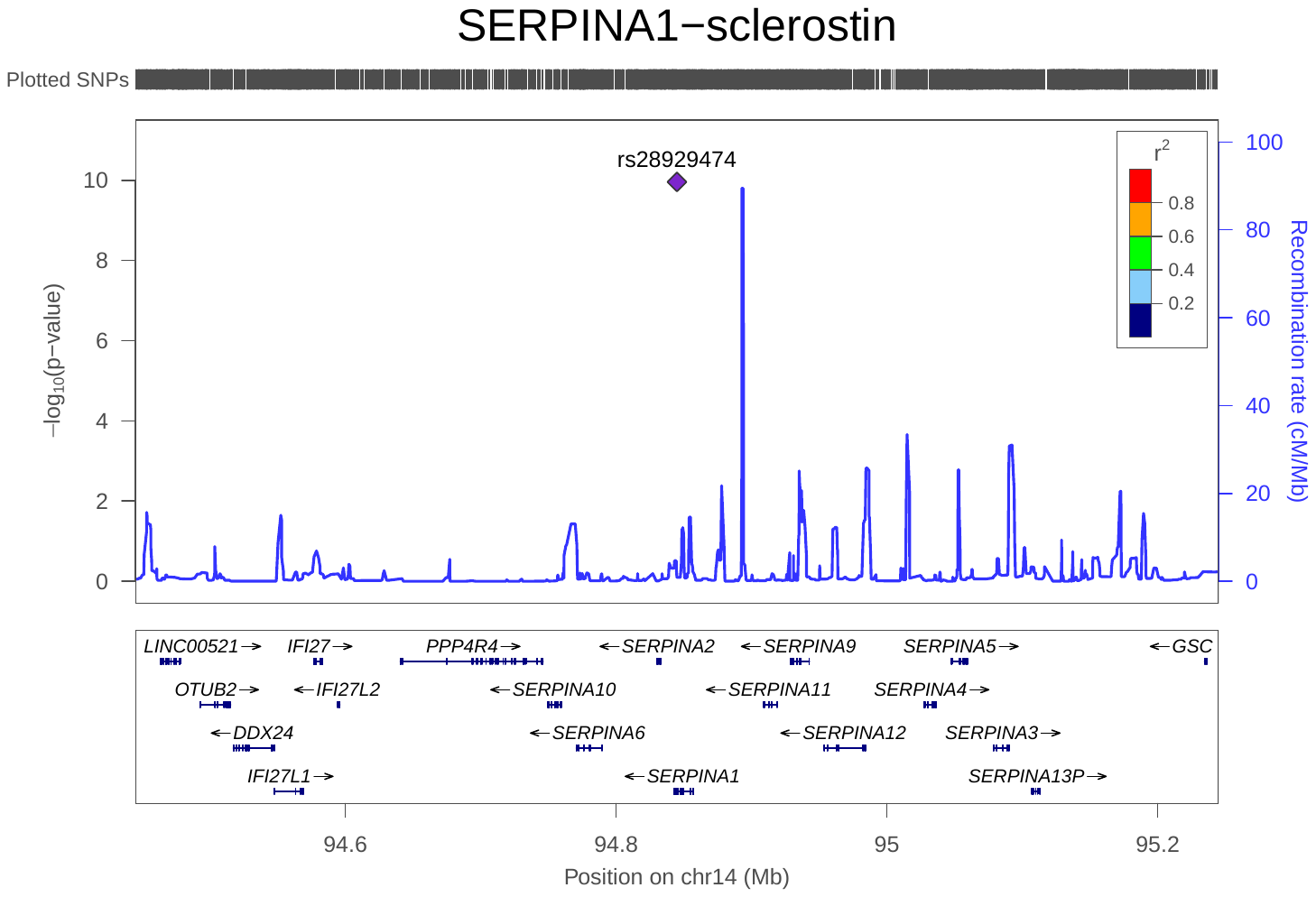

**Supplementary Figure 3**. Regional plot of gene expression level of *SERPINA1* and circulating sclerostin in the *SERPINA1* region. The two plots showed clearly that the genetic association peaks of the top and bottom plots did not overlap in this region.

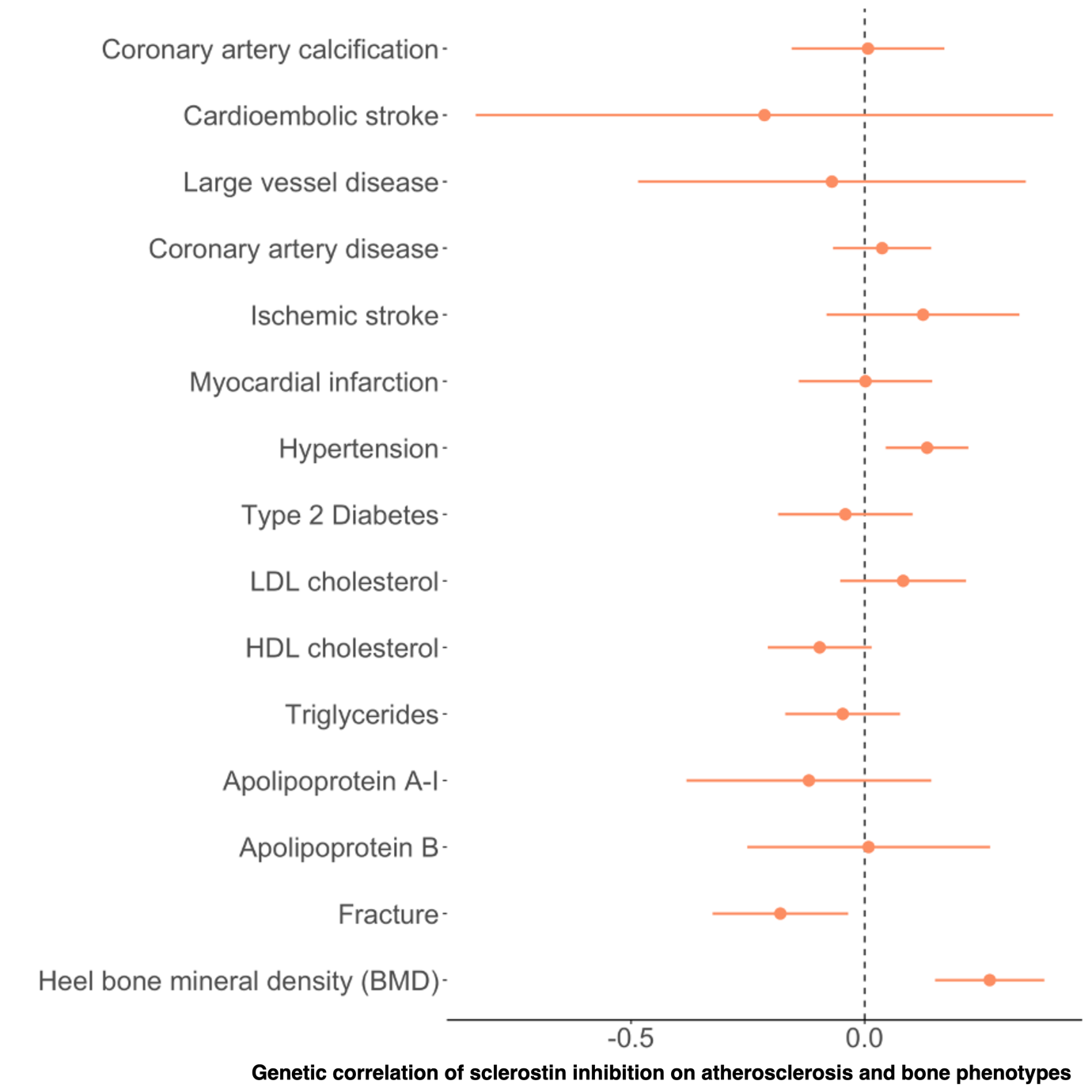

**Supplementary Figure 4**. Genetic correlation between sclerostin inhibition and atherosclerosis related and bone phenotypes.

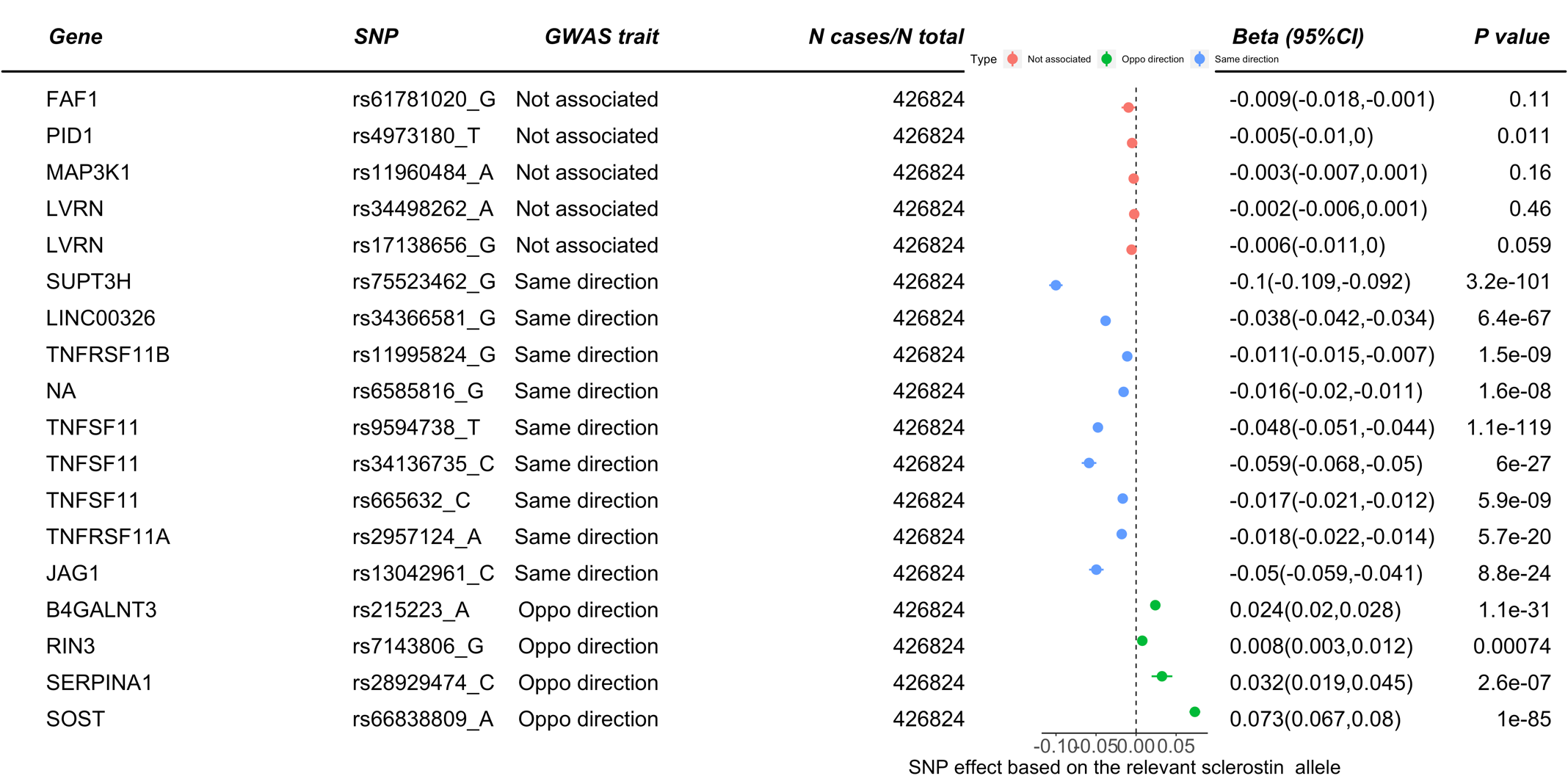

**Supplementary Figure 5**. Genetic effects of 18 sclerostin variants on eBMD. The alleles presented in the plot are the sclerostin-relevant alleles. Different colour refers to three types of variants (i) directionally similar effects on eBMD and sclerostin, (ii) directionally opposite effects on eBMD and sclerostin, and (iii) little effect of eBMD. Only variants with opposite direction were used as instruments for the Mendelian randomization analysis.

**Supplementary Note 1. Cohort Details of the nine studies involved in this GWAS meta-analysis**

The Avon Longitudinal Study of Parents and Children (ALSPAC)

ALSPAC is a prospective birth cohort which recruited pregnant women with expected delivery dates between April 1991 and December 1992 from Bristol UK. The initial number of pregnancies enrolled is 14,541 (for these at least one questionnaire has been returned or a “Children in Focus” clinic had been attended by 19/07/1999). Of these initial pregnancies, there was a total of 14,676 foetuses, resulting in 14,062 live births and 13,988 children who were alive at 1 year of age. Detailed information on health and development of children and their parents were collected from regular clinic visits and completion of questionnaires ^12^. Ethical approval was obtained from the ALSPAC Law and Ethics Committee and the Local Ethics Committees. Please note that the study website contains details of all the data that is available through a fully searchable data dictionary (http://www.bristol.ac.uk/alspac/researchers/our-data/).

Die Deutsche Diabetes Dialyse Studie (4D)

The 4D study was a prospective randomized controlled trial including patients with type 2 diabetes mellitus who had been treated by hemodialysis for less than 2 years ^3^. Between March 1998 and October 2002, 1255 patients were recruited in 178 dialysis centres in Germany. Patients were randomly assigned to double-blinded treatment with either 20 mg of atorvastatin (n = 619) or placebo (n = 636) once daily and were followed up until the date of death, censoring, or end of the study in March 2004. The primary end point of the 4D study was defined as a composite of death due to cardiac causes, stroke, and myocardial infarction, whichever occurred first. 4D study end points were centrally adjudicated by 3 members of the endpoints committee blinded to study treatment and according to predefined criteria. The study was approved by the medical ethics committees, and written informed consent was obtained from all participants.

The Gothenburg Osteoporosis and Obesity Determinants (GOOD)

The GOOD study was initiated to determine both environmental and genetic factors involved in the regulation of bone and fat mass ^4^. Male study subjects were randomly identified in the greater Gothenburg area in Sweden using national population registers, contacted by telephone, and invited to participate. To be enrolled in the GOOD study, subjects had to be between 18 and 20 years of age. There were no other exclusion criteria, and 49% of the study candidates agreed to participate (n = 1068). The study was approved by the ethics committee at the University of Gothenburg. Written and oral informed consent was obtained from all study participants.

MANOLIS Cohort

HELIC-MANOLIS (Minoan isolates) collection comprises individuals from the mountainous Mylopotamos villages, including Anogia, Zoniana, Livadia and Gonies (estimated population size of 6,000 in total) on the Greek island of Crete. It is one of the two cohorts composing the Hellenic Isolated Cohorts study (HELIC - http://www.helic.org). The specific population genetics ^5^ and dietary and lifestyle habits ^6^ of this cohort have been previously studied in the literature. The HELIC collections include blood for DNA extraction, laboratory-based haematological and biochemical measurements, and interview-based questionnaire data. The study was approved by the Harokopio University Bioethics Committee, and informed consent was obtained from human subjects. The MANOLIS cohort is named in honour of Manolis Giannakakis, 1978-2010

Fenland

The Fenland study is a population-based cohort of 12,435 participants of Caucasian-ancestry born between 1950 and 1975 who underwent detailed phenotyping at the baseline visit from 2005 to 2015. Participants were recruited from general practice surgeries in the Cambridgeshire region in the UK. Exclusion criteria were: clinically diagnosed diabetes mellitus, inability to walk unaided, terminal illness, clinically diagnosed psychotic disorder, pregnancy, or lactation. The study was approved by the Cambridge Local Research Ethics Committee (NRES Committee – East of England Cambridge Central, ref. 04/Q0108/19) and all participants provided written informed consent. The human biological samples were sourced ethically, and their research use was in accord with the terms of the informed consents under an IRB/EC approved protocol. Sclerostin levels (on the log2 scale) measured using the SomaLogic V4 assay were transformed using the rank-based inverse normal transformation^7^.

INTERVAL

The INTERVAL study comprises about 50,000 participants nested within a randomized trial of varying blood donation intervals^8^. Between mid-2012 and mid-2014, blood donors aged 18 years and older were recruited at 25 centres of England’s National Health Service Blood and Transplant (NHSBT). All participants gave informed consent before joining the study and the National Research Ethics Service approved this study (11/EE/0538). Participants completed an online questionnaire including questions about demographic characteristics (for example, age, sex, ethnicity), anthropometry (height, weight), lifestyle (for example, alcohol and tobacco consumption) and diet. Participants were generally in good health because blood donation criteria exclude people with a history of major diseases (such as myocardial infarction, stroke, cancer, HIV, and hepatitis B or C) and those who have had recent illness or infection. For SomaLogic assays, Sun et al randomly selected two non-overlapping subcohorts of 2,731 and 831 participants from INTERVAL. After genetic quality control, 3,301 participants (2,481 and 820 in the two subcohorts) remained for analysis. No statistical methods were used to determine sample size. The experiments were not randomized. Laboratory staff conducting proteomic assays were blinded to the genotypes of participants.

HUNT

The HUNT Study (Trøndelag Health Study) is a series of general health surveys of the adult population of the demographically stable Trøndelag county, Norway^9,10,11^ So far four health surveys have been conducted, HUNT1 (1984–1986), HUNT2 (1995–1997), HUNT3 (2006–2008) and HUNT4 (2017-2019). Approximately 230,000 individuals (aged ≥ 20 years) have participated in one or more HUNT surveys. More than 88,000 of these participants have been genotyped using one of the four Illumina HumanCoreExome arrays: 12 v.1.0, 12 v.1.1 and 24 with custom content (UM HUNT Biobank v1.0 and UM HUNT Biobank v2.0). Proteomic profiling was performed for 3532 subjects (cases with cardiovascular events and healthy controls) randomly selected from HUNT3 cohort (n=50,807). Out of these subjects, 1631 cases (both incident and prevalent) and 1297 healthy controls had complete genotyped data and thus were used for this study. The GWAS for protein was performed using rank transformed residuals adjusted for age, sex, batch effect, phase effect and principal components from 1-20. All individuals provided informed written consent and the study was approved by the Regional Committee for Medical and Health Research Ethics (REK # 2019/17355).

The Osteoarthritis Initiative (OAI)

The OAI is a prospective longitudinal study designed to identify risk factors for the incidence and progression of symptomatic tibiofemoral knee OA. A total of 4,796 men and women of any race/ethnicity aged 45 – 79 years were enrolled into pre-defined progression or incidence subcohorts^12^. Briefly, the progression subcohort included individuals who had symptomatic radiographic knee OA while the incidence subcohort included individuals who were at increased risk for developing symptomatic radiographic knee OA based on weight, knee symptoms, history of knee injuries/surgeries, family history of knee replacement and hand OA. Participants were recruited at four different clinical sites: 1) Brown University (Providence, RI); 2) The Ohio State University (Columbus, OH); 3) University of Maryland and The Johns Hopkins University (Baltimore, MD); and 4) University of Pittsburgh (Pittsburgh, PA). Details of the study protocol, including recruitment procedures and eligibility criteria are available on the OAI web site (<https://www.niams.nih.gov/grants-funding/funded-research/osteoarthritis-initiative>).

The OAI was genotyped on the Illumina Omni-Quad 2.5M array at the Translational Genomics Research Institute (Phoenix, AZ). Genotypes for were called using the Illumina BeadStudio software. The total number of genotyped SNPs was 2,440,283. Samples that had call rates across all SNPs of <95% were removed (185 samples). We additionally excluded from analysis potentially problematic samples based on 1) apparent mismatches between self-reported and genetically determined gender or 2) detection of second degree or higher relationships with other samples (56 samples). An additional 34 samples were excluded in whom we detected large chromosomal abnormalities using Log R Ratio (LRR) and B Allele Frequency (BAF), as described by others^13,14,15^. Genotypes for were imputed to the 1000 genomes CEU reference panel (June 2011 release) using the Minimac software program (<http://genome.sph.umich.edu/wiki/Minimac)>, resulting in a total of 8,248,570 imputed SNPs available for analyses after removal of SNPs with low minor allele frequencies (<1%) and poor imputation quality scores (<0.3).

Sclerostin was assessed in serum/plasma collected at the baseline visit using a sclerostin chemiluminescent assay (Medical University of Graz, Clinical Institute of Medical and Chemical Laboratory Diagnostics). The intra-assay coefficient of variation (CV) is <0.025 and the inter-assay CV is <0.05^16^. The assay can measure concentrations of 50-6500pg/ml, with a sensitivity (limit of quantitation) of <50pg/ml. The assay is specific for intact sclerostin.

The Ludwigshafen Risk and Cardiovascular Health (LURIC)

The Ludwigshafen Risk and Cardiovascular Health (LURIC) study is a prospective cohort study of individuals with and without cardiovascular disease and was designed to investigate environmental and genetic risk factors for the development of cardiovascular diseases. Between July 1997 and January 2000, 3316 participants of German ancestry were enrolled in the cardiology unit of a tertiary care medical centre in south-western Germany. The inclusion criteria were defined as clinical stability except for acute coronary syndromes (ACS), German ancestry, and availability of a coronary angiogram (indicated after standard clinical test diagnoses like chest pain and a positive, non-invasive stress test). Exclusion criteria were pre-specified as any acute illness other than ACS, any chronic disease where non-cardiac disease predominated, and a history of malignancy within the past five years. The detailed study protocol has been published^17^. Written informed consent was obtained from each participant prior to inclusion. The study was in accordance with the Declaration of Helsinki and approved by the ethics committee at the Medical Association of Rhineland-Palatinate (Ärztekammer Rheinland-Pfalz). Genotyping (Affymetrix 6.0 platform) and genotype calling (Birdseed v2 algorithm) were conducted at the LURIC study non-profit LLC, Heidelberg. Quality control and imputation to the 1000 Genomes phase I reference panel were performed as previously described^18^. After quality control, sclerostin measurements were available for n=1882 unrelated individuals.

**References**

1.	Boyd, A. *et al.* Cohort Profile: the ’children of the 90s’--the index offspring of the Avon Longitudinal Study of Parents and Children. *Int. J. Epidemiol.* **42**, 111–127 (2013).

2.	Fraser, A. *et al.* Cohort Profile: the Avon Longitudinal Study of Parents and Children: ALSPAC mothers cohort. *Int. J. Epidemiol.* **42**, 97–110 (2013).

3.	Wanner, C. *et al.* Randomized controlled trial on the efficacy and safety of atorvastatin in patients with type 2 diabetes on hemodialysis (4D study): demographic and baseline characteristics. *Kidney Blood Press. Res.* **27**, 259–266 (2004).

4.	Lorentzon, M., Swanson, C., Andersson, N., Mellström, D. & Ohlsson, C. Free testosterone is a positive, whereas free estradiol is a negative, predictor of cortical bone size in young Swedish men: the GOOD study. *J. Bone Miner. Res.* **20**, 1334–1341 (2005).

5.	Panoutsopoulou, K. *et al.* Genetic characterization of Greek population isolates reveals strong genetic drift at missense and trait-associated variants. *Nat. Commun.* **5**, 5345 (2014).

6.	Farmaki, A.-E. *et al.* The mountainous Cretan dietary patterns and their relationship with cardiovascular risk factors: the Hellenic Isolated Cohorts MANOLIS study. *Public Health Nutr.* **20**, 1063–1074 (2017).

7.	Pietzner, M. *et al.* Mapping the proteo-genomic convergence of human diseases. *Science* eabj1541 (2021) doi:10.1126/science.abj1541.

8.	Di Angelantonio, E. *et al.* Efficiency and safety of varying the frequency of whole blood donation (INTERVAL): a randomised trial of 45 000 donors. *Lancet* **390**, 2360–2371 (2017).

9.	Krokstad, S. *et al.* Cohort Profile: the HUNT Study, Norway. *Int. J. Epidemiol.* **42**, 968–977 (2013).

10.	Åsvold, B. O. *et al.* Cohort profile update: The HUNT study, Norway. *bioRxiv* (2021) doi:10.1101/2021.10.12.21264858.

11.	Brumpton, B. M. *et al.* The HUNT Study: a population-based cohort for genetic research. *bioRxiv* (2021) doi:10.1101/2021.12.23.21268305.

12.	Lester, G. The Osteoarthritis Initiative: A NIH Public-Private Partnership. *HSS J.* **8**, 62–63 (2012).

13.	Conlin, L. K. *et al.* Mechanisms of mosaicism, chimerism and uniparental disomy identified by single nucleotide polymorphism array analysis. *Hum. Mol. Genet.* **19**, 1263–1275 (2010).

14.	Peiffer, D. A. *et al.* High-resolution genomic profiling of chromosomal aberrations using Infinium whole-genome genotyping. *Genome Res.* **16**, 1136–1148 (2006).

15.	Laurie, C. C. *et al.* Quality control and quality assurance in genotypic data for genome-wide association studies. *Genet. Epidemiol.* **34**, 591–602 (2010).

16.	Drake, M. T. & Khosla, S. Hormonal and systemic regulation of sclerostin. *Bone* **96**, 8–17 (2017).

17.	Winkelmann, B. R. *et al.* Rationale and design of the LURIC study--a resource for functional genomics, pharmacogenomics and long-term prognosis of cardiovascular disease. *Pharmacogenomics* **2**, S1-73 (2001).

18.	Andlauer, T. F. M. *et al.* Novel multiple sclerosis susceptibility loci implicated in epigenetic regulation. *Sci Adv* **2**, e1501678 (2016).
